# Mycobiome Dysbiosis and Genetic Predisposition for Elevated IL-17A Drive Fibrosis in MASLD

**DOI:** 10.1101/2024.10.21.24315902

**Authors:** Nadja Thielemann, Sara Leal Siliceo, Monika Rau, Annika Schöninger, Nathalie Reus, Alexander M. Aldejohann, Aia Shehata, Isabell S. Behr, Natalie E. Nieuwenhuizen, Michaela Herz, Heike M. Hermanns, Mohammad Mirhakkak, Jürgen Löffler, Thomas Dandekar, Kerstin Hünniger-Ast, Ronny Martin, Gianni Panagiotou, Andreas Geier, Oliver Kurzai

## Abstract

Metabolic dysfunction-associated steatotic liver disease (MASLD) is the leading cause of chronic liver disease in Western countries. Progression to metabolic dysfunction-associated steatohepatitis (MASH) occurs when fat accumulation in the liver triggers Th17 activation and other inflammatory processes. In this study, we identify the *IL17A* rs2275913 minor allele variant as a risk factor for fibrosis progression in MASLD patients. In patients with advanced fibrosis, we also observed an increased abundance of fungal CTG species including *Candida albicans* and *Debaryomyces hansenii*, which are potent triggers of Th17 responses. Integrating genetic risk-predisposition and mycobiome composition, we show in *ex vivo* T cell stimulation assays, that donors carrying the minor allele variant of *IL17A* rs2275913 secreted significantly higher IL-17A levels in response to CTG species. Additionally, MASH patients carrying the *IL17A* rs2275913 risk allele have elevated Th17/Treg ratios in peripheral blood. Taken together, our data indicate that genetic predisposition for enhanced Th17 responses in the context of mycobiome dysbiosis can trigger MASH progression and liver fibrosis.

**Graphical Abstract:** 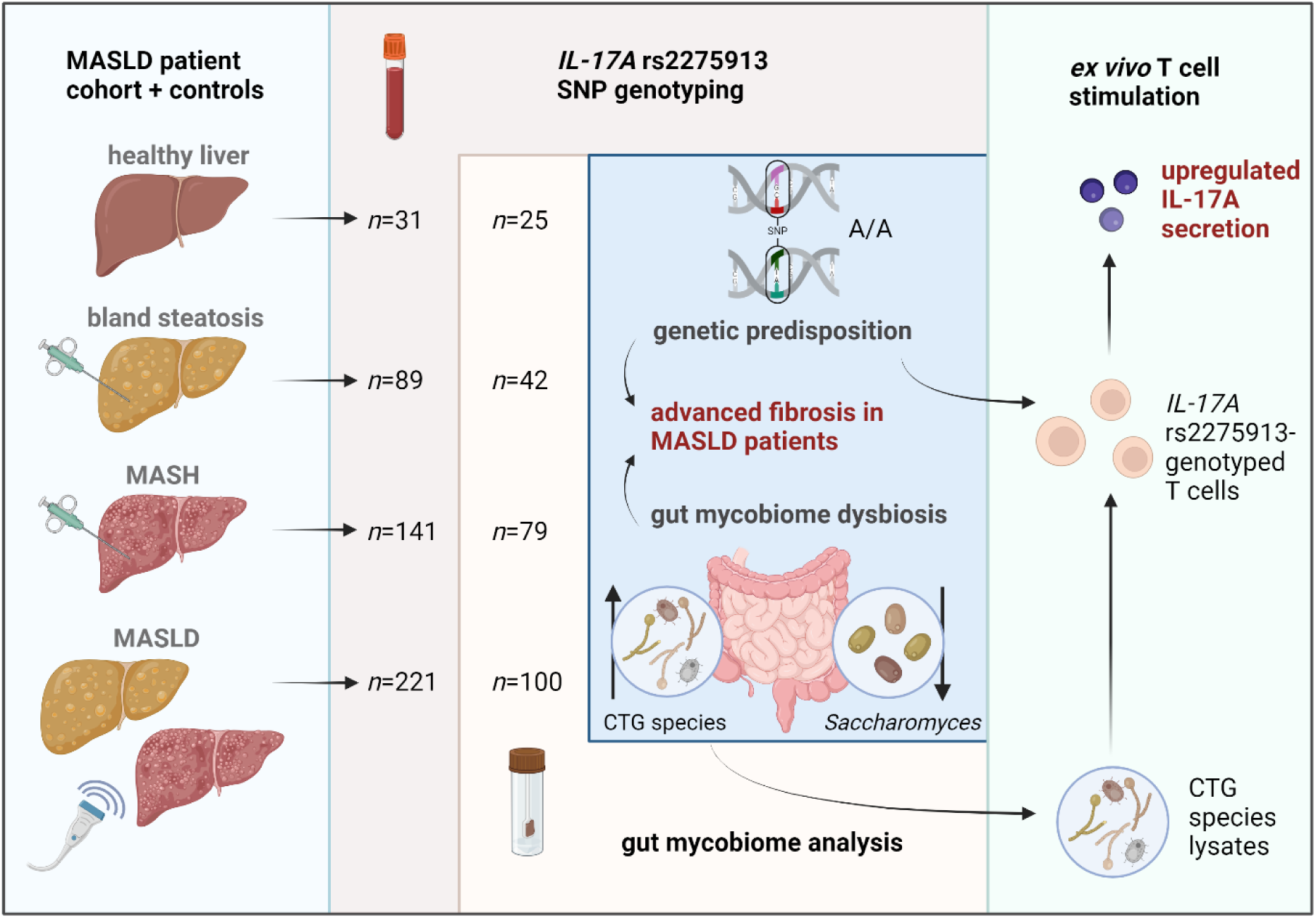

This Graphical Abstract was created with BioRender.com.

**Brief summary:** Increased antifungal immune responses triggered by gut mycobiome dysbiosis in genetically predisposed patients can lead to severe stages of metabolic dysfunction-associated steatotic liver disease.

## INTRODUCTION

Metabolic dysfunction-associated steatotic liver disease (MASLD, formerly known as non-alcoholic fatty liver disease [NAFLD, (1)]) has become one of the leading causes of chronic liver diseases, with a global prevalence of approximately 25% (2). MASLD is characterized by the accumulation of excess fat in the liver in the absence of relevant alcohol consumption and commonly associated with obesity, type 2 diabetes and metabolic syndrome (3). Fat accumulation in hepatocytes leads to bland steatosis (BS), the initial step of MASLD pathogenesis (4). Continued fat accumulation and lipotoxicity trigger inflammation and the transition to metabolic dysfunction-associated steatohepatitis (MASH). These inflammatory processes lead to the development of fibrosis which may ultimately progress to cirrhosis (5). The reasons why some patients progress to MASH and others do not remain unclear, though evidence suggests that exaggerated Th17 responses are associated with progression to MASH (6).

The liver receives approximately 75% of its blood supply via the portal vein and therefore has close connection to the human intestinal tract, which is densely colonized by microorganisms collectively known as the microbiome (7). Gut microbiota dysbiosis has been repeatedly observed in obesity and type 2 diabetes mellitus (8, 9). Recent data provide clear evidence that the composition of gut microbiota also has a direct impact on MASLD pathogenesis (10–13). Previously, we identified a significantly higher abundance of short chain fatty acid (SCFA)-producing bacteria such as *Fusobacteriaceae*, *Prevotellaceae*, and *Ruminococcaceae* in the gut of patients with advanced MASLD (14).

In contrast to the bacterial microflora, the role of intestinal fungi in MASLD is less well understood. Mycobiome research faces several challenges, including non-standardized protocols, technical difficulties and incomplete reference databases (15, 16). A key challenge is the unresolved taxonomy of the polyphyletic genus *Candida*, which comprises relevant human gut mycobionts that are only distantly related. Within the genus *Candida*, the CTG species, which translate the CTG codon predominantly to serine instead of leucine, are of special importance in human gut colonization. CTG species comprise both opportunistic pathogens (e.g. *Candida albicans*) and non-pathogenic species (e.g. *Debaryomyces hansenii*) (17). *C. albicans* is a major human mycobiont and has been shown to be a direct inducer of human antifungal Th17 cell responses (18). Cross-reactivity to *C. albicans* induces Th17 responses to other fungi and plays an important role in airway inflammation, especially during acute allergic bronchopulmonary aspergillosis. Thus, immune activation triggered by *C. albicans* may represent a key mechanism driving Th17 responses with broad systemic impact (18).

Recently, Demir *et al.* characterized a distinct fecal mycobiome signature in non-obese MASLD patients with *Malassezia* spp. abundance increased in patients with BS and *C. albicans* and *Penicillium* spp. abundance elevated in MASH patients. Notably, increased intestinal *C. albicans* colonization was associated with increased levels of systemic antibodies against *C. albicans* and advanced fibrosis (19). Furthermore, the presence of *C. albicans* specific T-cells in the liver has been demonstrated in alcohol-associated liver disease (ALD) (20).

The aim of our study was to investigate the role of the gut mycobiota in MASH in the context of intrinsic variations in IL-17A signaling. Our results identify a novel *IL17A* genetic risk variant for liver fibrosis in MASH and show that intestinal colonization with *C. albicans* and related species (CTG species) can contribute to enhanced inflammation in the presence of this genotype.

## RESULTS

### Study population

A total of 482 European subjects were recruited for this study, including 230 histology-proven MASLD patients (89 BS and 141 MASH). **Fig. 1** illustrates the clinical and histological phenotypes of the study participants in a flow diagram. Stool samples could be collected from a sub-cohort of subjects (42 BS, 79 MASH, 100 MASLD). As an additional control, a group of healthy individuals (HC) was included. Patients with and without 6 months antibiotic-free intervals were analyzed separately.

**Fig. 1.**
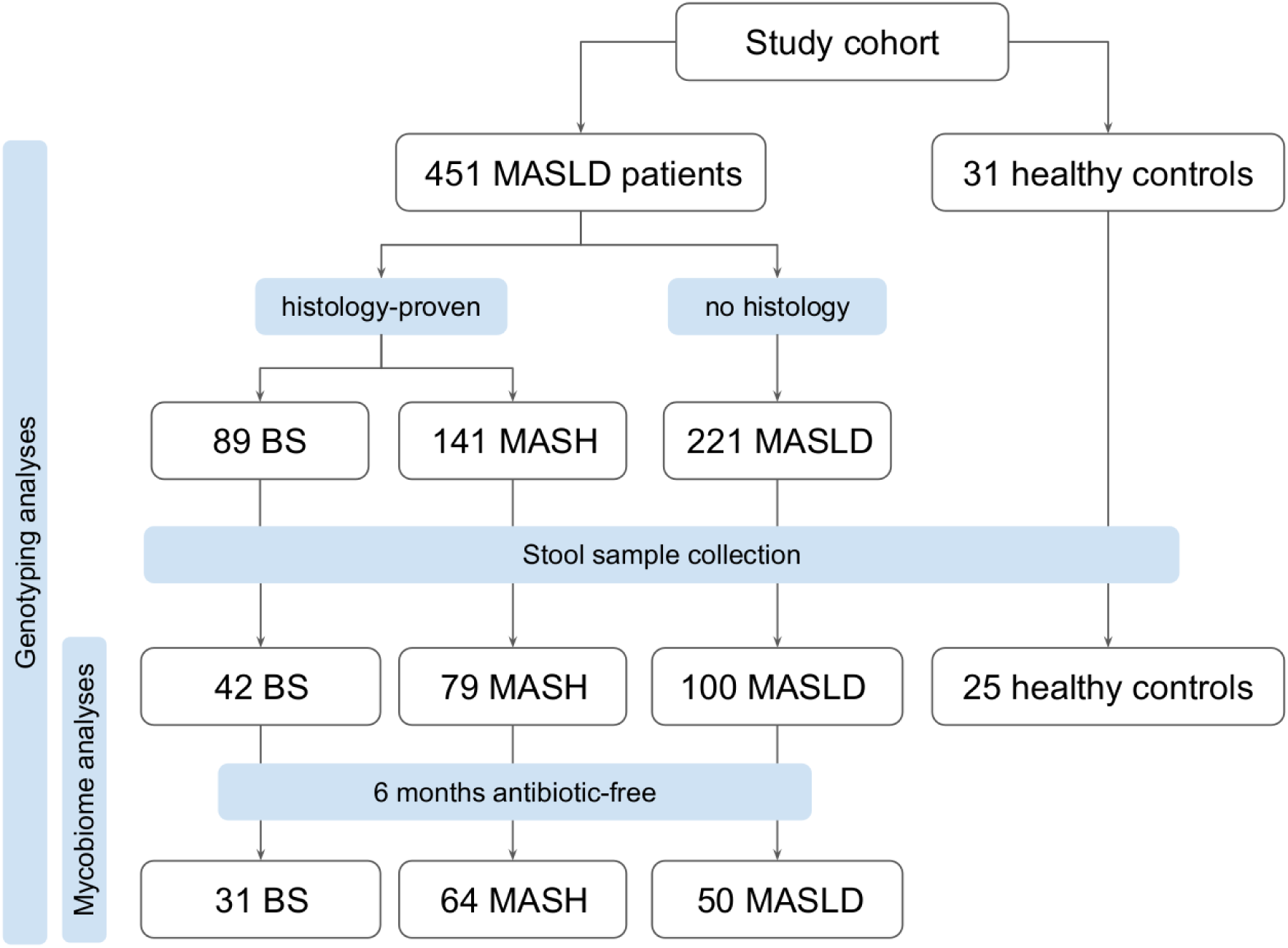
Flow diagram with an overview of the study participants.

### Genetic variation in *IL17A* predisposes patients to develop fibrotic MASLD

Th17 responses and IL-17A signaling are known to play a key role in MASH related inflammation (6). To further investigate genetic factors governing Th17 activation in our patient cohort, we focused on analyzing relevant SNPs. As a preliminary step, we first validated the association of the *PNPLA3* rs738409 genotype with liver fibrosis assessed by fibroscan values for the dataset available for our patient cohort (21) as it is a major genetic risk factor for MASLD (22) (p_Kruskal-Wallis_=0.105; p_glm_=0.028; glm adjusted; **Fig. 2A**). Subsequently, all consecutive SNP-based glm calculations for the SNPs analyzed in this study were adjusted for the *PNPLA3* rs738409 risk genotype. In an extensive dbSNP database search for genetic variants in Th17 signaling-associated genes linked to gastrointestinal disease with inflammatory properties, we identified three potential candidate SNPs. No significant association with MASLD disease parameters was found for rs16910526 (*CLEC7A* gene, coding for Dectin-1) and rs4077515 (*CARD9*) (**Suppl. Fig. 1**). Our third candidate SNP was the *IL17A* rs2275913 SNP. This SNP was previously shown to be associated with inflammatory bowel disease (23). TaqMan SNP genotyping of our 451-patient MASLD cohort identified 175 G/G (homozygous for major allele variant, 38.8%), 55 A/A (homozygous for minor allele variant, 12.2%), and 221 A/G (heterozygous, 49%) genotypes (**Fig. 2B**). Genotype frequencies were in Hardy-Weinberg equilibrium and selection for specific genotypes was excluded (**Suppl. Fig. 2**). The calculated minor allele frequency (MAF) of 36.7% is comparable to the published ALFA European cohort MAF of 34.85%. Statistical analysis revealed a significant association between the *IL17A* rs2275913 genotype and liver fibrosis assessed by fibroscan (p_Kruskal-Wallis_=0.368; p_glm_=0.029; generalized linear model (glm) adjusted; **Fig. 2C**). Patients carrying the minor allele (A/A & A/G) showed increased liver stiffness and more severe fibrosis compared to those with a homozygous major allele genotype (G/G).

**Fig. 2.**
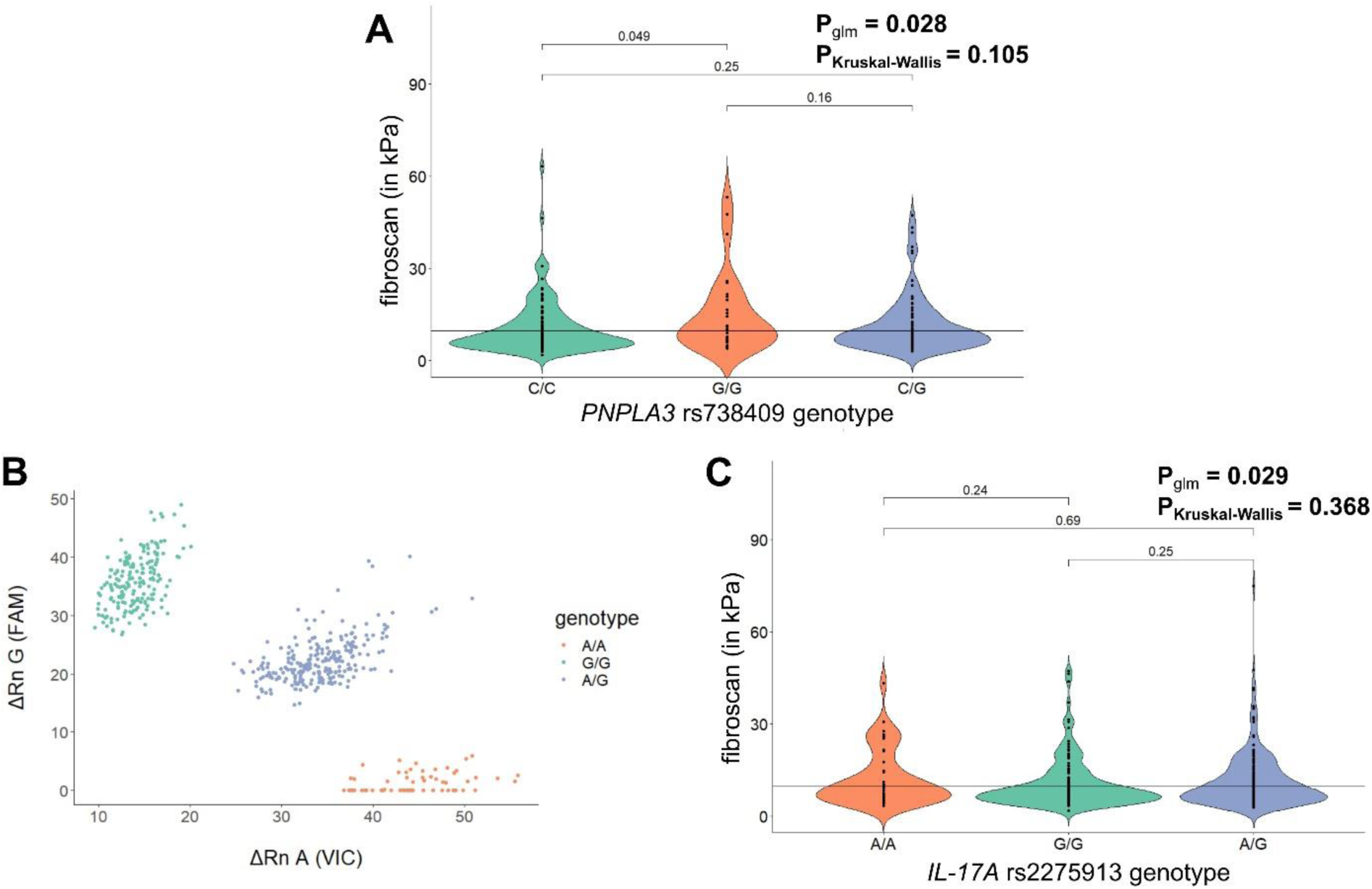
The *IL17A* rs2275913 genotype is associated with liver stiffness in MASLD. **A**) Violin Plot for visualization of *IL17A* genotype association with fibrosis as assessed by fibroscan. Statistical comparison was performed using Kruskal-Wallis Test (p_Kruskal-Wallis_) and generalized linear models adjusted for age, gender, BMI, *PNPLA3* rs738409 genotype (p_glm_) based on fibroscan cut-off=9.7 kPa. **B**) Allelic discrimination Plot after TaqMan SNP Genotyping. **C)** Violin Plot for visualization of known *PNPLA3* risk variant rs738409 association with fibrosis as assessed by fibroscan. Statistical comparison was performed using Kruskal-Wallis Test (p_Kruskal-Wallis_) and generalized linear models adjusted for age, gender and BMI (p_glm_) based on fibroscan cut-off=9.7kPa.

### A distinct mycobiome composition characterizes MASH patients

Intestinal colonization by *C. albicans* is a major inducer of Th17 responses (18) and prior data suggested altered mycobiome composition in subgroups of MASLD patients, potentially linking mycobiome dysbiosis to Th17 activation (19). Thus, we analyzed mycobiome composition using ITS1 libraries for 145 subjects from our study cohort to estimate the fungal genus and species abundance and explore the possible role of fungi in MASLD progression and liver damage. On average, we generated 15,500 high-quality, non-chimeric reads per sample and fungal annotation identified 29 genera and 223 species in total. Genus-level fungal profiles showed that *Saccharomyces*, *Penicillium*, and CTG species were the topmost abundant fungal colonizers among our study participants, at 16.7%, 16.1%, and 12.5%, respectively. We used the CTG species group for genus clustering, as *Candida* is a polyphyletic genus comprising a large variety of phylogenetically distant species. Members of the CTG species found in our dataset are pathogenic *C. albicans*, *Candida tropicalis*, *Candida dubliniensis*, *Candida parapsilosis* and non-pathogenic *D. hansenii.* Although still commonly referred to as *Candida,* other species including *Nakaseomyces glabratus* (*Candida glabrata), Kluyveromyces marxianus* (formerly *Candida kefyr*) and *Pichia kudriavzevii* (formerly *Candida krusei*) are only distantly related to the CTG species and were analyzed separately (17, 24).

In total, 76 BS, MASH, and MASLD subjects in our cohort reported antibiotic use within 6 months before the stool collection. As a recent study showed that antibiotics may have a long-term influence on the mycobiome (25) we investigated whether antibiotics had a noticeable impact on the gut mycobiome profiles of the different disease groups. We found that the mycobiome alpha-diversity measured by the Shannon and Simpson index at genus level was significantly increased in MASH subjects who had used antibiotics compared to the antibiotic-free subjects (Wilcoxon rank-sum test, p_Shannon index_=0.028 and p_Simpson index_=0.025; **Suppl. Fig. 3**). However, no differences were found in the BS and MASLD groups between antibiotic and antibiotic-free subjects. Using Aitchison distance to compute beta-diversity, no differences were found between the antibiotic and antibiotic-free subjects in any of the disease groups (PERMANOVA adjusted for age, gender and obesity-related parameters, p>0.05). Nevertheless, to avoid a possible impact of antibiotics on the downstream analysis, two approaches were used for the mycobiome comparisons. For all the main results, unless specified, a dataset with only the long-term antibiotic-free samples was used. Alternatively, mycobiome analysis was performed using all samples, adjusting for antibiotic intake when appropriate (see methods for details).

To study the mycobiome changes related to MASLD progression, we first performed pairwise comparisons between BS, MASH, MASLD and HC in alpha diversity measured by the Shannon and Simpson index and we found no significant differences between the four diagnosed groups (Wilcoxon rank-sum test, p>0.05 for all pair group comparisons for Shannon and Simpson index, data not shown). Beta diversity analysis using Aitchison distance to assess the overall mycobiome community differences showed that the fungal composition was significantly different between MASH and HC subjects (PERMANOVA adjusted for age, gender and obesity-related parameters, p=0.01, **Fig. 3A**).

**Fig. 3.**
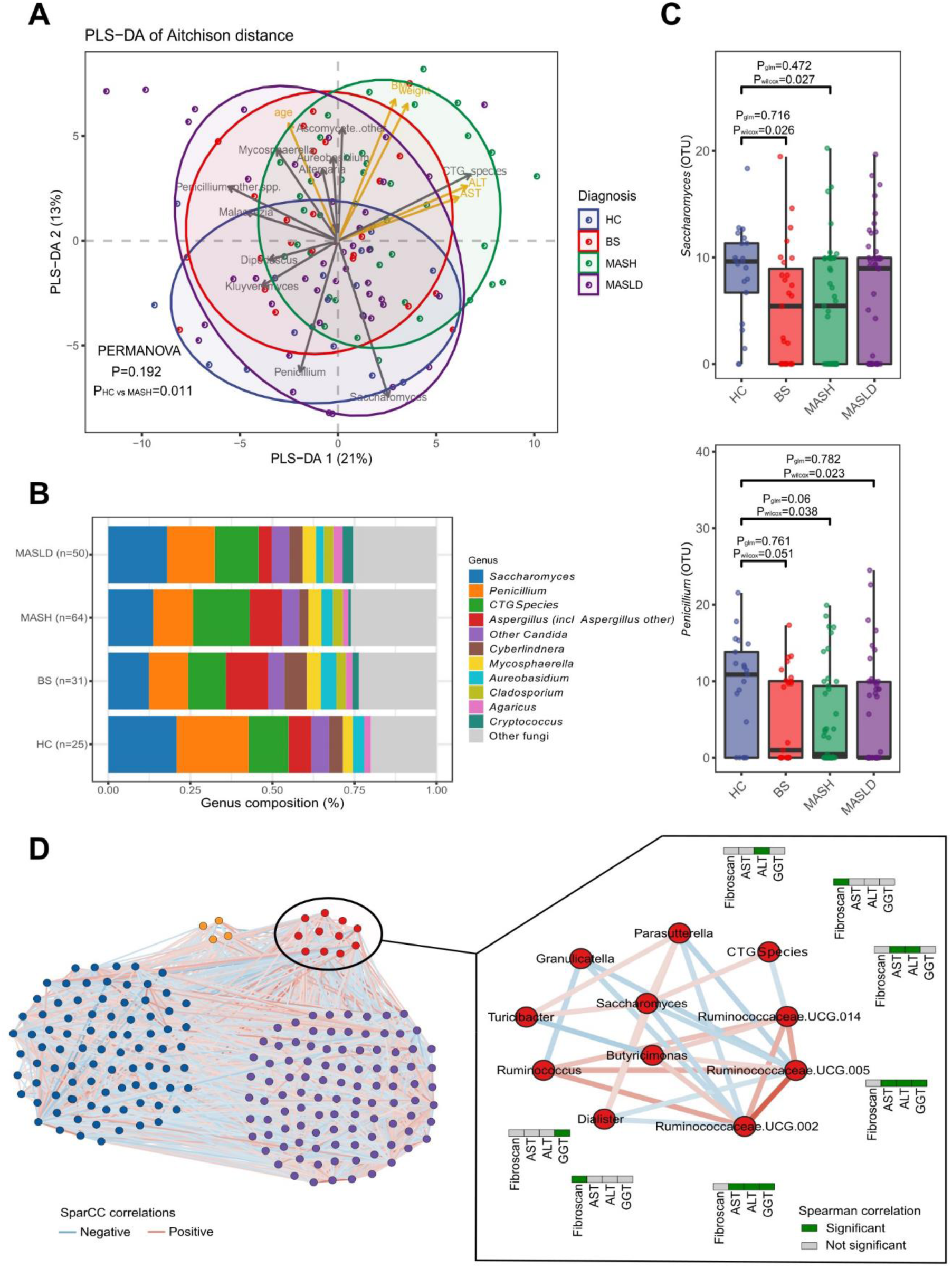
Mycobiome changes in the different diagnosed groups and healthy controls and microbial community network. **A**) Beta diversity. PLS-DA of Aitchison distance of the mycobiome composition by diagnosis. **B**) Overview of mycobiome composition at genus level in MASLD, BS, MASH, and HC groups. **C**) Boxplot of *Saccharomyces* and *Penicillium* abundances. Statistical comparison between groups (HC, BS, MASH, and MASLD) was performed using Wilcoxon rank-sum test (p_wilcoxon_) and generalized linear models adjusting for age, gender and obesity-related parameters (p_glm_). **D**) Microbial community network showing the 4 subcommunity modules. Significant negative correlations are shown in blue and positive in red. The module significantly associated with MASLD-related parameters is shown with red nodes and significant correlations between the genera and fibroscan, AST, ALT, and GGT are shown in green.

We then explored the differences in fungal abundance between the disease groups (BS, *n*=31; MASH, *n*=64; MASLD, *n*=50) and healthy controls (HC, *n*=25). Again, fungi were grouped according to genus, except for the CTG species group. The most abundant genus in the HC group was *Penicillium* (22.2%), followed by *Saccharomyces* (20.9%), and CTG species (12.2%) (**Fig. 3B**). Similar abundance patterns were observed for the BS and MASLD groups (**Fig. 3B**). Interestingly, in MASH, the most abundant genus was the CTG species group (17.8%), followed by *Saccharomyces* (14.1%) and *Penicillium* (12.5%) (**Fig. 3B**). Among the most abundant genera, we found *Saccharomyces* abundance significantly decreased in BS and MASH in comparison to HC (Wilcoxon rank-sum test, p_HCvsBS_=0.026, p_HCvsMASH_=0.027, **Fig. 3C**). In addition, *Penicillium* was significantly decreased in abundance in all disease stages in comparison to HC even though it did not reach statistical significance in comparison to BS (Wilcoxon rank-sum test, p_HCvsMASH_=0.038, p_HCvsMASLD_=0.023, p_HCvsBS_=0.051, **Fig. 3C**). However, the statistical significance was lost for the two genera when accounting for age, gender, and obesity-related parameters (glm adjusted, p>0.05, **Fig. 3C**), suggesting a potential confounding effect in the abundances of the two genera by these factors.

We subsequently repeated all the analytical steps using the full cohort and not only the antibiotic-free subjects and confirmed the significant differences in beta diversity between the MASH and HC groups (PERMANOVA adjusted, p=0.034, **Suppl. Fig. 4A**) and the significant decrease in abundance of *Penicillium* in the MASLD and MASH groups compared to HC (p_HCvsMASH_=0.03, p_HCvsMASLD_=0.02, Wilcoxon rank-sum test; p_HCvsMASH_=0.007, p_HCvsMASLD_=0.42, glm adjusted). Even though it did not reach statistical significance as it did when using the antibiotic-free set of samples, the same trend was observed for *Saccharomyces,* having lower abundance in the MASH and MASLD groups compared to HC (Wilcoxon rank-sum test, p_HCvsMASH_ and p_HCvsMASLD_ < 0.1).

Finally, we used 16S data from our cohort in order to build a microbial community network to identify possible associations between fungal and bacterial genera and MASLD progression. Using all cohort samples, we built a community network using FastSpar (26), and identified a total of 5,848 significant correlations (SparCC, p<0.05) from which 4,017 remained significant after multiple testing correction (FDR correction, q<0.1). Using greedy modularity optimization, a total of 4 subcommunities were identified in the full network (**Fig. 3D**). We then studied the associations between these subcommunity modules and MASLD and identified one module that consists of 2 fungal (CTG species group and *Saccharomyces*) and 9 bacterial genera (including *Ruminococcus*, *Dialister*, and *Parasutterella* amongst others) that were significantly associated with MASLD-related parameters (fibroscan, AST, ALT, and GGT) (Fisher’s Exact test, p=0.049, odds ratio=3.580), suggesting the interplay of the two microbial kingdoms and MASLD.

### Increased abundance of CTG species in patients with advanced fibrosis

To investigate whether changes in the mycobiome composition are linked to progression of liver fibrosis we classified the subjects into early or advanced fibrosis groups using a fibroscan cut-off value of 9.7kPa (27). Alpha diversity analysis (Shannon and Simpson index) revealed no significant differences at the genus level (Wilcoxon rank-sum test, p>0.05). However, beta diversity analysis using Aitchison distance showed significant differences in the mycobiome composition between early and advanced fibrosis groups (PERMANOVA adjusted for age, gender, and obesity-related parameters, p=0.007, **Fig. 4A**). Further exploration of the mycobiome composition (**Fig. 4B**) revealed that CTG species were significantly increased in the advanced compared to the early fibrosis group even when accounting for age, gender, and obesity-related parameters (p_wilcoxon_=0.0009, Wilcoxon rank-sum test; p_glm_=0.002, glm adjusted, **Fig. 4C**).

**Fig. 4.**
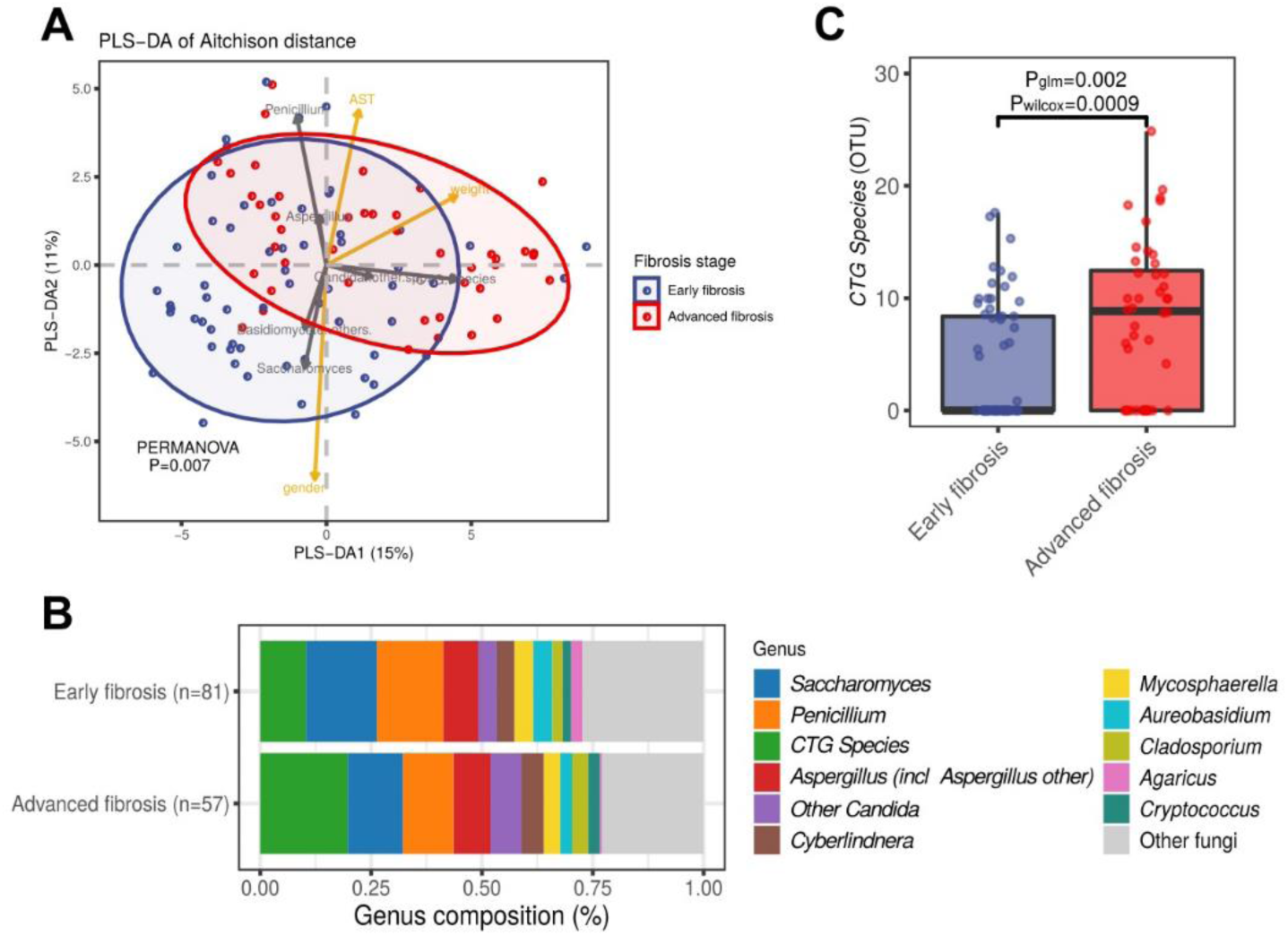
Mycobiome changes by fibroscan-based fibrosis groups. **A**) Beta diversity. PLS-DA of Aitchison distance of the mycobiome composition by fibrosis stage group. **B**) Overview of mycobiome composition at genus level in early and advanced fibrosis groups (fibroscan cut-off </> 9.7 kPa). **C**) Boxplot of CTG species abundances. Statistical comparison between early and advanced fibrosis was performed using Wilcoxon rank-sum test (p_wilcoxon_) and generalized linear models adjusting for age, gender and obesity-related parameters (p_glm_).

To further corroborate our findings, we calculated the beta diversity (Aitchison distance) of early and advanced fibrosis groups using all subjects and not only the antibiotic-free subjects. Again, significant differences were identified (PERMANOVA adjusted, p=0.01, **Suppl. Fig. 4B**) and a significant increase in CTG species abundance in advanced-compared to early fibrosis (Wilcoxon rank-sum test, p_wilcoxon_=0.0007; glm adjusted, p_glm_=0.002) was also clearly visible when analyzing the full cohort (**Suppl. Fig. 4C**). Thus, in both antibiotic-free and total study cohorts, CTG species abundance is significantly higher in the advanced fibrosis group, suggesting that these species may contribute to disease progression.

Regression analysis between fibroscan stiffness values independent of the arbitrary cut-off of 9.7kPA and CTG species abundance also revealed a significant association (glm adjusted for age, gender, and obesity-related parameters, p=0.001, estimate=0.22). Correlation analysis also showed a positive significant correlation between fibroscan values and CTG species abundances (Spearman’s correlation adjusted, p=0.026, ρ=0.23). We also evaluated this association for the complete cohort and the same results were obtained (presence/absence of CTG species associated with the fibrosis stage, p=0.002, odds ratio=2.73, Fisher’s Exact test; CTG species abundances and fibroscan, significant positive correlation, Spearman’s correlation adjusted, p=0.01, ρ=0.20 and glm adjusted, p=0.002, estimate=0.23, accounting for age, gender, obesity-related parameters and antibiotic use).

Finally, the trend of increasing CTG species abundance was also visible when samples were grouped by fibrosis stage obtained by histology (**Suppl. Fig. 5**), although for fibrosis stages F3 and F4 the sample size was too small to reach statistical significance due to a lack of biopsied patients (Kruskal-Wallis, p=0.07 for the antibiotic-free sample set and p=0.086 for the full cohort). We further explored CTG species imbalance in an advanced fibrosis stage in the antibiotic-free set of samples and found an association between the presence/absence of the CTG species group and the fibrosis stage (Fisher’s Exact test, p=0.006, odds ratio=3.097).

### CTG species trigger increased pro-inflammatory responses in the presence of the *IL17A* rs2275913 risk genotype

Our findings so far establish a new risk genotype in *IL17A* for progression of MASLD and confirm mycobiome dysbiosis in patients with advanced stage liver fibrosis. To determine whether the *IL17A* rs2275913 SNP has an impact on responses to CTG species, we stimulated freshly isolated T cells from rs2275913-genotyped donors *ex vivo* with fungal lysates. To ensure that differences in T cell proportions among peripheral blood mononuclear cells (PBMCs) of individual donors did not influence IL-17A levels, we first isolated T cells and then used equal numbers of T cells in the *ex vivo* stimulation assays. An age-dependent influence on CD4+ T cell frequency was excluded due to similar mean age of donors within the genotype groups (mean values for donor age are between 30-31 years). T cells were stimulated with fungal lysates of a pathogenic (*C. albicans*) and a non-pathogenic (*D. hansenii*) representative of the CTG species group as well as with the non-CTG species *Saccharomyces cerevisiae*. Resulting levels of multiple Th17 signaling-associated cytokines were quantified using Luminex technology. T cell functionality was measured after stimulation with anti-CD3/anti-CD28 and did not show genotype-dependent differences validating all effects induced by fungal lysates as species-specific. To account for medium-mediated activation effects, all samples were normalized to the corresponding medium control values for each donor. Both CTG species lysates triggered increased release of proinflammatory IFN-γ, TNF-α, IL-22 and IL-17A following *ex vivo* T cell stimulation especially in donors carrying the *IL17A* rs2275913 A allele (**Table 1**). Cytokine levels were generally lower after T cell stimulation with the non-CTG species *S. cerevisiae* in comparison to *C. albicans* and *D. hansenii* (**Table 1**). To further characterize especially proinflammatory IL-17A release following *ex vivo* T cell stimulation of rs2275913-genotyped donors, we additionally measured IL-17A release by highly sensitive ELISA after calculation with a 4-parameter standard fit curve. Again, effective and rs2275913 genotype-independent T cell functionality was evaluated after stimulation with anti-CD3 (**Suppl. Fig. 6A**), and all samples were normalized to the corresponding medium control values for each donor. T cells were stimulated with fungal lysates of pathogenic (*C. albicans*) and non-pathogenic (*D. hansenii*) representatives of the CTG species group (28, 29), a combination of both stimuli as well as with the non-CTG species *S. cerevisiae* (**Fig. 5**). Both CTG species lysates induced IL-17A secretion and T cells from individuals with the rs2275913 A/A genotype showed significantly increased IL-17A levels in comparison to those with the rs2275913 G/G and heterozygous genotypes (*C. albicans*: p_Kruskal-Wallis_=0.104, p_A/AvsG/G_ =0.042; *D. hansenii*: p_Kruskal-Wallis_=0.065, p_A/AvsG/G_ = 0.035; **Fig. 5A+B**). Interestingly, this effect was even elevated when both fungal lysates were used as stimuli for T cells in half concentration each (p_Kruskal-Wallis_=0.019, p_A/AvsG/G_ = 0.019; **Fig. 5C**), indicating a cumulative effect of antigens of these two species as seen before (18). Additionally, the strongly elevated IL-17A secretion in donors with the rs2275913 A/A variant were not visible after T cell stimulation with non-CTG species *S. cerevisiae* (**Fig. 5D**) or pathogenic *C. tropicalis*, *C. parapsilosis* and *N. glabratus* (**Suppl. Fig. 6B-D**). Thus, the *IL17A* rs2275913 genotype modifies the amount of IL-17A produced in response to specific CTG species. Together with the elevated CTG species abundance in MASLD patients, this suggests a combinatory effect of a genetically determined enhanced Th17 response and the imbalance in CTG species on fibrosis progression in patients with MASH.

**Fig. 5.**
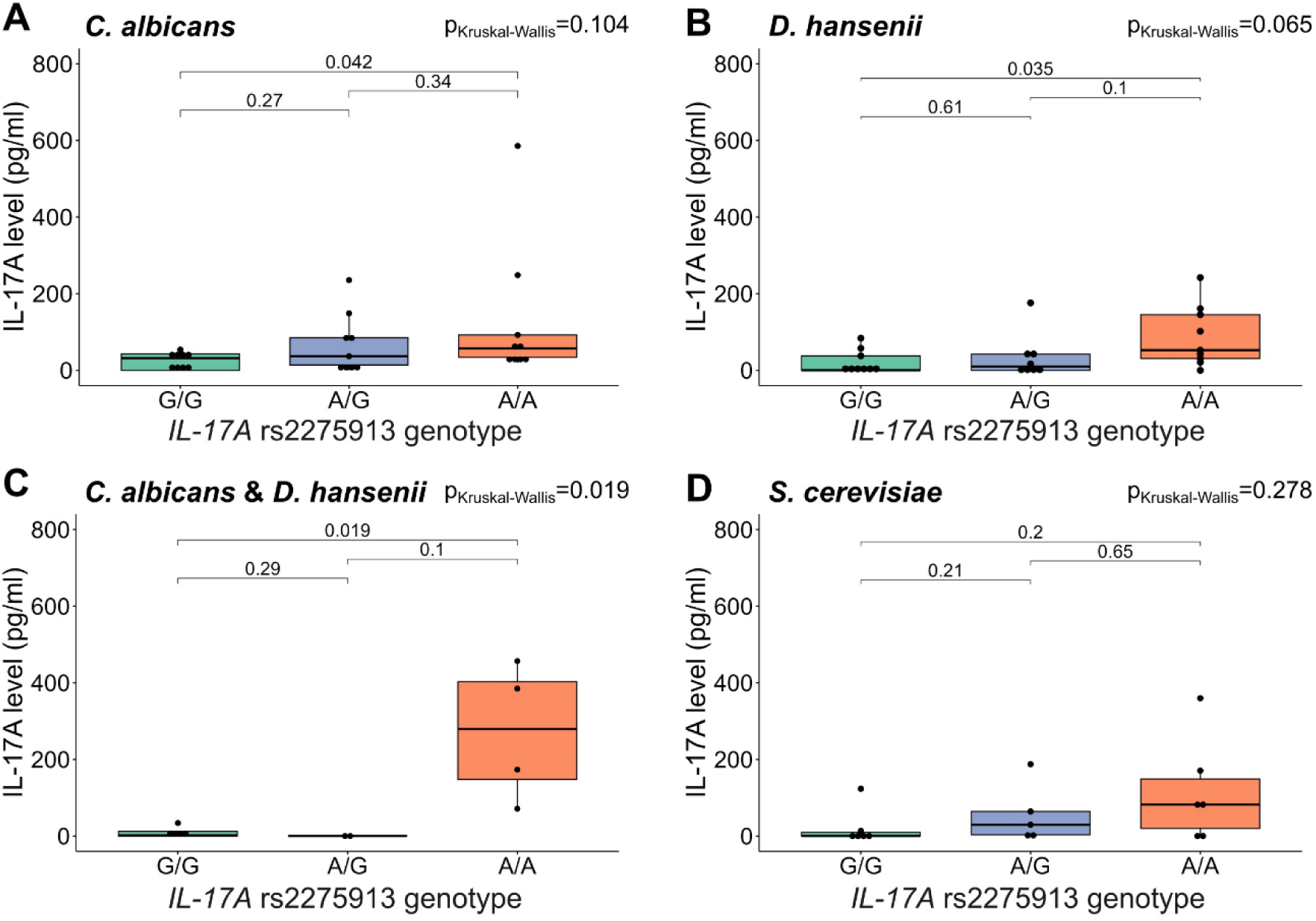
Increased IL-17A production in T cells from subjects homozygous for the rs2275913 minor allele variant. T cells were stimulated with fungal lysates and IL-17A concentrations in samples were measured by ELISA and calculated with a 4-parameter standard fit curve. 27 subjects were included in this assay. Due to interindividual variation of T cell numbers not all stimuli were tested for each condition. IL-17A secretion after stimulation with **A**) *C. albicans* lysate (G/G: *n=9*, A/G: *n=9*, A/A: *n=9*), **B**) *D. hansenii* lysate (G/G: *n=9*, A/G: *n=8*, A/A: *n=9*), **C)** *C. albicans* and *D. hansenii* lysate (G/G: *n=5*, A/G: *n=2*, A/A: *n=4*) and **D**) *S. cerevisiae* lysate (G/G: *n=6*, A/G: *n=5*, A/A: *n=6*). Statistical comparisons for **A**-**D** were performed using Kruskal-Wallis Test (p_Kruskal-Wallis_) and T test comparing mean IL-17A values between genotypes. Horizontal lines in the boxplots indicate from top to bottom 75th percentile, median and 25th percentile. Whiskers display minimum and maximum values in 1.5x the interquartile range. Dots specify individuals for the three *IL17A* rs2275913 genotypes.

**Table 1.** Cytokine levels after e*x vivo* stimulation of T cells from *IL17A* rs2275913-genotyped donors with *C. albicans*, *D. hansenii & S. cerevisiae*. Values are shown as means and range.

|  | *C. albicans* |  |  | *D. hansenii* |  |  | *S. cerevisiae* |  |  |
| --- | --- | --- | --- | --- | --- | --- | --- | --- | --- |
|  | G/G<br>*n* = 9 | A allele<br>carrier<br>*n* = 18 | p | G/G<br>*n* = 9 | A allele<br>carrier<br>*n* = 17 | p | G/G<br>*n* = 6 | A allele<br>carrier<br>*n* = 11 | p |
| **IFN-γ** | 240.3<br>(0-1661) | 1411.0<br>(0-4825) | 0.199 | 169.3<br>(0-717.2) | 1902.9<br>(0-7833) | 0.019 | 46.8<br>(0-211.2) | 1290.8<br>(0-6155.3) | 0.339 |
| **IL-17A** | 17.1<br>(0-49.0) | 100.2<br>(0-680.9) | 0.169 | 10.1<br>(0-45.7) | 98.8<br>(0-635.6) | 0.057 | 3.4<br>(0-20.2) | 66.2<br>(0-315.0) | 0.264 |
| **IL-22** | 52.1<br>(0-185.7) | 253.5<br>(0-1275) | 0.066 | 22.2<br>(0-95.8) | 206.4<br>(0-1038) | 0.137 | 4.5<br>(21.3) | 126.3<br>(0-648.2) | 0.174 |
| **TNF-α** | 29.7<br>(0-79.2) | 181.0<br>(0-861.1) | 0.127 | 1.6<br>(0-7.2) | 94.2<br>(0-405.7) | 0.028 | 2.4<br>(0-11.4) | 48.8<br>(0-282.9) | 0.465 |

### MASH patients carrying the *IL17A* rs2275913 A allele have elevated Th17/rTreg-ratios

To further explore the potential predisposition of MASLD patients to develop MASH if they carry the *IL17A* rs2275913 minor allele, we associated results from *IL17A* rs2275913 genotyping of MASLD patients to their blood Th17/resting regulatory T cells (rTreg) ratios. In a previous work, we identified elevated Th17/rTreg-ratios especially in MASH patients in comparison to healthy controls (6), which was also observed in the subset of patients enrolled in this study (p_MASHvsHC_=0.00012, p_Kruskal-Wallis_=0.00016; **Fig. 6A**). *IL17A* rs2275913 genotyping of these patients revealed that Th17/rTreg-ratios are significantly upregulated in carriers of the A allele (p_A/GvsG/G_=0.033, p_Kruskal-Wallis_=0.066; **Fig. 6B**). Taken together with the results from *ex vivo* stimulation assays, this indicates that MASLD patients may be predisposed to develop MASH if they carry the *IL17A* rs2275913 A allele due to contribution of increased Th17/rTreg-ratios as well as increased pro-inflammatory cytokine release to liver inflammation, particularly in the presence of a dysbiotic mycobiome characterized by increased abundance of CTG species.

**Fig. 6.**
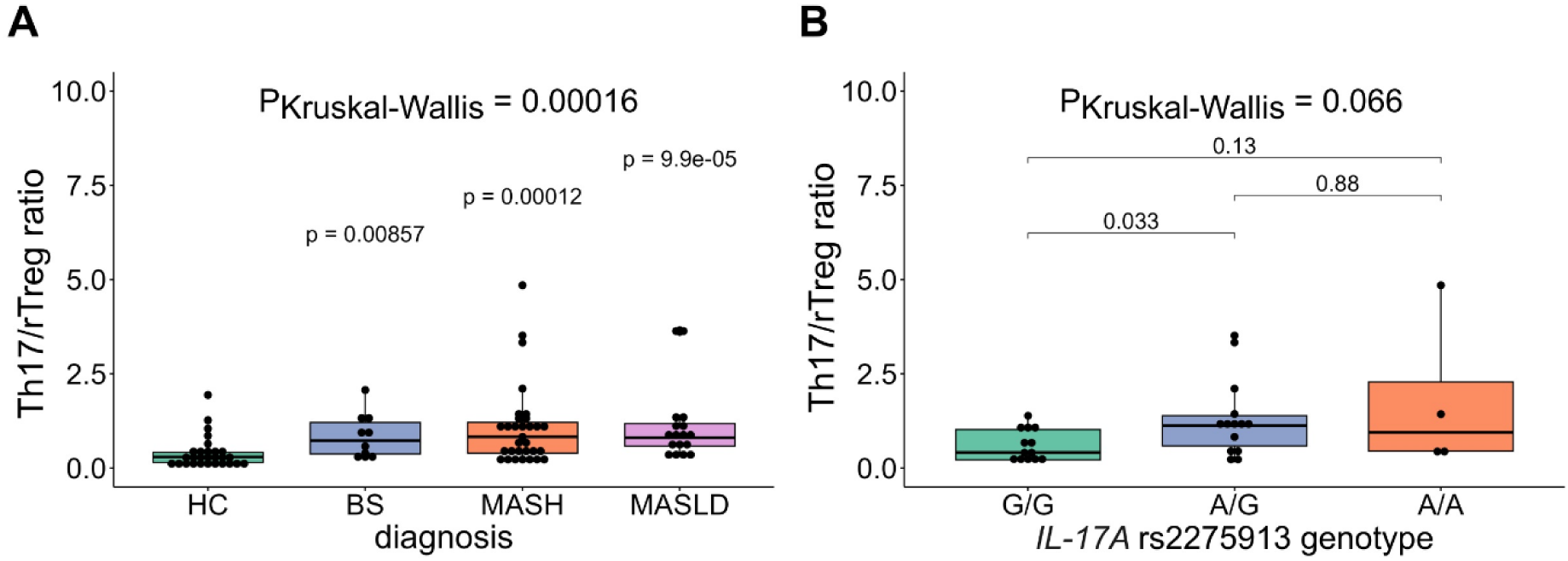
Elevated Th17/rTreg ratios in MASH patients carrying the *IL17A r*s2275913 minor allele. **A)** Th17/rTreg ratios in blood samples of healthy controls and MASLD patients included in this study (HC: *n=28*, BS: *n=10*, MASH: *n=31* and MASLD: *n=17*). **B)** Th17/rTreg ratios in MASH patients according to *IL17A* rs2275913 genotype (G/G: *n=13*, A/G: *n=14*, A/A: *n=4*). Statistical comparisons were performed using Kruskal-Wallis Test (P_Kruskal-Wallis_) and T test comparing mean Th17/rTreg ratios using HC as a reference group (**A**) and between *IL17A* rs2275913 genotypes (**B**). Horizontal lines in the boxplots indicate from top to bottom 75th percentile, median and 25th percentile. Whiskers display minimum and maximum values in 1.5x the interquartile range. Dots specify individuals for the three *IL17A* rs2275913 genotypes.

## DISCUSSION

MASLD, formerly known as NAFLD, affects more approximately 25% of the global population and as such constitutes a major public health challenge world-wide. Its pathogenesis is multifactorial, involving multiple determinants like genetic predisposition, diet and composition of the microbiota (30). In this study, we identify a link between genetic variation in *IL17A* and the dysbiosis of the gut mycobiome as contributing factors to MASLD progressions, particularly inflammation-dependent progression to MASH. Our previous research showed that the progression from BS to MASH is marked by an increase in IL-17A-producing cells within intrahepatic CD4+ T cells and a higher Th17/rTreg ratio in peripheral blood (6). Here, we found that patients with the *IL17A* rs2275913 minor allele (A/A genotype) were at a higher risk of severe fibrosis, displayed elevated IL-17A secretion in response to fungal stimuli, and had higher Th17/rTreg ratios.

The involvement of IL-17A in maintaining health during immune responses to infection, injuries and physiological stress is well described (31). Importantly, IL-17A is also crucial for the antifungal response of the adaptive immune system (32). However, if dysregulated, this proinflammatory cytokine can contribute to the development of various diseases including liver fibrosis (33). Dysbiosis of the intestinal mycobiome correlates with inflammatory bowel disease as well as liver diseases (19, 34, 35). Our observations indicate that both, genetic variation in *IL17A* and increased intestinal abundance of CTG species, may have a combined effect to foster the progression of BS to MASH. This is in line with recently published work characterizing the contribution of Th17 cells, which are specific to *C. albicans*, to ALD indicating similar mechanisms for MASLD pathogenesis and especially MASH development (20). On the other hand, increased *C. albicans* commensal gut colonization positively correlates with systemic levels of fungal-specific Th17-driven inflammation and therefore might improve host defense against other pathogens (36). These observations clearly support the importance of a balanced antifungal IL-17A-mediated immunity for human health and disease that seem to be dysregulated in *IL17A* rs2275913 A/A predisposed MASLD patients and contributes to inflammatory-driven liver fibrosis.

The described increase of intestinal CTG species abundance in MASLD patients, especially in those with advanced fibrosis, is similar to the elevated abundance of *C. albicans* and *Debaryomyces* spp. found in the gut of alcohol abuse disorder patients (35). The *C. albicans*-secreted exotoxin candidalysin is associated with the severity of liver disease in ALD patients (37). Additionally, it was found that *C. albicans* strain diversity influences the immune response in inflammatory bowel disease (38). Candidalysin might explain how highly abundant intestinal *C. albicans* contribute to gastrointestinal and liver diseases. However, the gene encoding this peptide is absent in most non-*albicans* CTG species, including *D. hansenii* (28). Our experiments revealed that *D. hansenii* can similarly trigger IL-17A secretion following *ex vivo* T cell stimulation, resulting in comparable cytokine levels as observed for *C. albicans*. This finding indicates different immune recognition mechanisms for the several CTG species that could culminate in comparable activation patterns of Th17 responses. This might be similar to the different mechanisms of immune recognition discussed for closely related but non-CTG species such as *N. glabratus* (39). Irrespective of the way of activation, elevated CTG species-mediated IL-17A secretion may contribute to BS to MASH transition and is probably enhanced in carriers of the *IL17A* rs2275913 A/A variant.

Mycobiome dysbiosis involving increased intestinal abundance of *D. hansenii* is of particular interest, as this food-borne yeast is often found on cheese as well as processed meat in Western-style diet and is therefore often seen as a transient mycobiome component. Although the possible probiotic properties of *D. hansenii* have been studied intensively (40), its functional role in the context of human disease is still unknown and needs to be characterized. Here, *D. hansenii* was shown to possess a T cell stimulatory potential, that is enhanced in *IL17A* rs2275913 A/A-predisposed MASLD patients. Prior studies established that *D. hansenii* contributes to impaired wound healing in Crohńs disease patients as well as in the corresponding mouse model (41). If mice with elevated intestinal abundance of *C. albicans* and / or *D. hansenii* underwent antifungal amphotericin B treatment, symptoms of impaired wound healing and also diet-induced liver fibrosis and steatohepatitis improved (19, 41). Overall, these data combined with our observations suggest that non-pathogenic CTG species are relevant for gut- and liver-related inflammatory diseases, especially in genetically predisposed *IL17A* rs2275913 A/A patients.

Although our results corroborate the role of intestinal fungi, especially CTG species, and antifungal Th17 responses in MASLD pathogenesis, there are some limitations. Our ITS1-based gut mycobiome analysis clearly confirmed recent data generated by ITS2 sequencing. However, we cannot exclude that primer bias resulted in the omission of common mycobiome-associated species like *Malassezia* spp. in our data set (42). Due to the intestinal mycobiome variability between and even within individuals, future longitudinal studies are essential to exclude possible diet, antibiotic or environmental-mediated effects. These studies would further reinforce causal intestinal mycobiome changes associated with MASLD pathogenesis. Incorporating the bacterial influence on the interaction between CTG species and antifungal Th17 responses in MASLD pathogenesis would be of interest for future studies. Although the analysis of this triangle including human-fungal-bacterial interaction might be challenging under *in vitro* conditions, our interaction analysis already predicted that CTG species and SCFA-producing bacterial genera jointly contribute to liver pathology and MASLD progression. The bacterial-derived SCFAs might act as soluble mediators in this interactome as they were already linked to both, MASLD pathogenesis and increased intestinal abundance of *C. albicans*, before (43, 44).

Altogether, our results provide deeper insights into the role of intestinal fungi in MASLD pathogenesis as they suggest a combinatory effect of *IL17A* risk variant-driven increased antifungal Th17 response and elevated intestinal CTG species abundance to drive liver inflammation and progression to fibrosis, thereby promoting BS to MASH transition.

## METHODS

### Patients (MASLD cohort)

In this prospective study, 451 MASLD patients were enrolled between 2016-2019 in the Division of Hepatology of the Department of Medicine II, University Hospital Würzburg, Germany. All study participants were >18 years old and diagnosed with MASLD by histology (n=230) and/or clinically by transient elastography (TE; fibroscan and controlled attenuation parameter (CAP) (n= 350). We included all clinically characterized MASLD subjects in our cohort irrespective of histological characterization to investigate associations between genetic variations in antifungal immunity and gut mycobiome imbalance with the largest possible sample size. Although liver histology is considered the gold standard of MASLD diagnosis, the more easily accessible TE is a widely used and validated technique that has shown a high performance for the diagnosis and exclusion of advanced fibrosis when compared to liver biopsy (45). Additionally, it reduces the imminent risk of sampling error due to heterogeneous distribution of fibrosis when assessing liver biopsy specimens (46).

Clinical and anthropometric characteristics of the study cohort are shown in **Table 2**. A cutoff for daily alcohol consumption was set (<20 g/d for female and <30 g/d for male subjects) and further underlying liver disease (e.g., autoimmune liver disease or chronic viral hepatitis) was excluded. Information on patient’s last antibiotic treatment was documented. Fecal, serum and whole blood samples were immediately snap-frozen and stored in the local biobank.

**Table 2.** MASLD patient cohort characteristics. Values are shown as means and range.

|  | **MASLD patients**<br>(*n*=451) | **healthy controls**<br>(*n*=31) |
| --- | --- | --- |
| **general information** |  |  |
| *male* | *n*=166 (37.5%) | *n*=15 (48.4%) |
| *female* | *n*=277 (62.5%) | *n*=16 (51.6%) |
| *age (years)* | 46.5 (18-73), *n*=451 | 27.3 (23-37), *n*=31 |
| *BMI (kg/m<sup>2</sup>)* | 46.2 (21.6-78.2), *n*=450 | 21.4 (17.5-30), *n*=31 |
| *underweight: (<18.5)* | *n*=0 | *n*=4 (12.9%) |
| *normal: (18.5-24.9)* | *n*=9 (2%) | *n*=23 (74.2%) |
| *overweight: (25-29.9)* | *n*=45 (10%) | *n*=3 (9.7%) |
| *obese – type I: (30-34.9)* | *n*=31 (7%) | *n*=1 (3.2%) |
| *obese – type II: (35-39.9)* | *n*=29 (6.5%) | *n*=0 |
| *obese – type III: (>40)* | *n*=336 (74.5%) | *n*=0 |
| **liver function tests** |  |  |
| *AST (U/L)* | 36.8 (11-249), *n*=450 | 20.5 (11.6-45.6), *n*=27 |
| *ALT (U/L)* | 49.4 (5.8-469.7), *n*=451 | 18.5 (10-46.6), *n*=28 |
| *γ-GT (U/L)* | 65.2 (7.6-914), *n*=450 | *NA* |
| *AP (U/L)* | 77.2 (0-222), *n*=450 | *NA* |
| *AST/ALT ratio* | 0.9 (0.2-3.7), *n*=450 | 1.2 (0.6-1.6), *n*=27 |
| *glucose (mg/dl)* | 111.2 (70-444), *n*=430 | *NA* |
| **lipid metabolism** |  |  |
| *cholesterol (mg/dl)* | 187.5 (22-342), *n*=419 | *NA* |
| *triglyceride (mg/dl)* | 166.8 (31-1188), *n*=419 | *NA* |
| **elastography** |  |  |
| *fibroscan (kPa)* | 11.6 (1.8-75), *n*=350 | *NA* |
| *CAP (dB/m)* | 346.5 (40-400), *n*=258 | *NA* |

### DNA extraction from blood and PBMCs and TaqMan SNP Genotyping

DNA was extracted from frozen blood or PBMC samples using the Roche High Pure PCR Template Preparation Kit (Sigma Aldrich, #11796828001) according to the manufacturer’s instructions. Isolated DNA was then used in TaqMan SNP Genotyping Assays (ThermoFisher, CN #4351376; CARD9 (ID: C 25956930_20), CLEC7A (ID: C 33748481_10), IL-17A rs2275913 (ID: C 15879983_10) according to manufacturer’s instructions. Assays were conducted with the qTower^3^ (Analytik Jena) and analyzed with the qPCRsoft 3.4 software (Analytik Jena). The functionality of TaqMan SNP Genotyping was confirmed by additional sequencing of 5% samples and validating the obtained genotypes. For sequencing, a 414 bp part of interest in the *IL17A* gene was amplified (5’: ATATGATGGGAACTTGAGTAGTTTCCG, 3’: CTCCTTCTGTGGTCACTTACGTGG) with 2x Q5 polymerase master mix according to the manufacturer’s instructions (NEB, #M0492L). PCR samples were purified with the PCR & Gel Clean-Up Kit (Macherey-Nagel, #740609.50) according to the manufacturer’s instructions and sent to LGC genomics for sequencing with the 5’ primer. DNA sequences were evaluated with ApE (v.3.0.8).

### Fecal DNA extraction, internal transcribed spacer 1 and 16S rRNA sequencing

Microbial DNA was extracted from stool samples using the DNeasy PowerSoil Kit (Qiagen, #12888-100) according to the manufactureŕs instructions. We divided the sample into 4 subsamples to increase efficiency of the beat-beating step.

The Illumina platform Miseq V3 with paired-end reads of 300 bp was used for all samples. For the ITS sequencing samples were processed by LGC Genomics GmbH. The ITS1 region was amplified using ITS1F/ITS2R primers. The total read count was on average 54,000 reads/sample. From the 246 total 16S rRNA sequencing samples, 149 16S rRNA sequencing samples that had not been previously analyzed were processed by LGC Genomics GmbH using sequencing primers 341F-785R, targeting the V3-V4 region. The total read count was on average 56,000 reads/sample. 97 16S rRNA sequencing samples from a previous study were processed as described in Rau et al (14).

### Taxonomic profiling

Taxonomic annotation of fungal Internal Transcribed Spacer (ITS) was performed using the PIPITS pipeline (47) version 2.4, with default parameters including quality filtering, read-pair merging, ITS1 extraction and chimera removal. Remaining reads were binned based on 97% similarity as operational taxonomic units (OTUs) and aligned with QIIME (48) to the UNITE fungi database (49) using mothur classifier. Samples were then normalized by cumulative sum scaling using the R package metagenomeSeq. Due to the complex fungal taxonomy, we grouped fungi according to genus but used the CTG species to characterize the *Candida* genus.

For the 16S rRNA sequencing data, quality control to remove low-quality reads and taxonomic annotation was performed using QIIME (48). Raw reads were joined and trimmed with cutadapt to remove the primer sequences. Deblur workflow was used for filtering and denoising the joined reads. Assigning taxonomic information to each amplicon sequence variant (ASV) was performed using a Naive Bayes classifier with 99% similarity in QIIME. The classifier was fitted to the appropriate rRNA gene region (V3-V4) with the SILVA 132 database (50).

### Diversity analysis

Alpha diversity indices detailing mycobiome community composition within samples were calculated using the R package vegan. Testing for significant differences in alpha diversity was performed using Wilcoxon rank-sum test. For estimating beta diversity reflecting community dissimilarities, *cmultRepl* function from R package zCompositions was first used to perform Bayesian-Multiplicative replacement of count zeros to the raw OTU table. Aitchison distances were calculated using *aDist* function from the R package robCompositions. We performed Partial Least Squares Discriminant Analysis (PLS-DA) using the mycobiome Aitchison distance matrix with the R package mixOmics. To test for significant differences in the mycobiome composition, permutational multivariate analysis of variance (PERMANOVA) as implemented in the function *adonis* from R package vegan adjusting for age, gender, obesity-related parameters (age, gender, BMI, DM, aHT and hyperlipidemia) was used. Mycobiome community and clinical data (age, gender, height, weight, BMI, AST and ALT) were fit onto the ordination using the function *envfit* from vegan R package.

### PBMC and T cell isolation

Freshly drawn blood from healthy volunteers was diluted 1:1 in PBS / 1 mM EDTA (Invitrogen, ThermoFisher Scientific, #AM9260G) containing 1% inactivated human AB serum (Sigma Aldrich, #H4522-100ML) and separated via Biocoll density gradient medium (Bio&SELL, #BS.L 6115) in SepMate tubes (Stemcell Technologies, #85460) according to the manufacturer’s instructions. Afterwards, PBMCs were washed 3 times with PBS-EDTA-human serum mix. As T cell proportions vary strongly between individual PMBC donors, we additionally isolated T cells before stimulation.

T cells were isolated from freshly isolated PBMCs by negative selection with the human Pan T Cell Isolation Kit (Miltenyi, #130-096-535) according to manufacturer’s instructions and the purity of >90% was assessed by flow cytometry (Miltenyi MACSQuant^Ⓡ^).

PBMC and T cell number and cell viability were measured directly after isolation with the LUNA automated cell counter (Logos Biosystems) and the viability was in each case >99%.

### Preparation of fungal lysates

50 ml inoculated YPD medium (20 g/L glucose, 20 g/L peptone, 10 g/L yeast extract) was cultured overnight at 25 °C (*D. hansenii* CBS767) and 37 °C (*C. albicans* SC5314, *N. glabratus* CBS138, *C. parapsilosis* ATCC22019, *C. tropicalis* PI941, *S. cerevisiae* AR#0400). The overnight culture was diluted 1:50 in 50 ml YPD medium and cultured for another 5 h. Cells were harvested by centrifugation at 4.000 g for 10 min and the cell pellet was resuspended in lysis buffer (50mM Tris-HCl, 150 mM NaCl, 0.1 % Triton X-100, 1 mM DTT, 10 % glycerol) with freshly adjusted proteinase inhibitor (Sigma, #S8820-20TAB). For lysis, 500 µl glass beads were added per tube and five 1min vortexing steps were followed by five 1 min cooling steps on ice. After centrifugation at 20.000 g for 5 min the supernatant was transferred to a fresh reaction tube and stored in aliquots at -80 °C. The protein concentration was measured with the Qubit protein assay kit (Invitrogen, ThermoFisher Scientific, #Q33211).

### *Ex vivo* T cell stimulation

Freshly isolated T cells were plated at 2×10^6^ cells/well in 48-well plates and stimulated with 40 µg/ml fungal lysate or CTL-Test^TM^ culture medium supplemented with PenStrep and L glutamine as medium control, in a final volume of 500 µl. For control of effective T cell functionality, wells were precoated with 1 µg/ml anti-human CD3 antibody (Miltenyi, #130-093-387) at 37 °C for 2 h before addition of cells and medium. All samples were supplemented with 1 µg/ml anti-human CD28 antibody (Miltenyi, #130-093-375). The plates were incubated for 48 h at 37 °C with 5 % CO2. All samples were prepared in duplicates. After incubation, supernatants were frozen at -80 °C until cytokine measurement.

### Quantification of cytokines by multiplex immunoassay

The secretion of cytokines (IL-17A, IFN-γ, IL-22, TNF-α) was assessed in supernatants of *ex vivo* T cell stimulation assays using Luminex technology (ProcartaPlex™ Multiplex Immunoassay, Thermo Fisher Scientific). The analyses were performed according to the instructions from the manufacturer.

### IL-17A ELISA

Antigen-specific IL-17A levels were measured in thawed supernatants in duplicate using the IL-17A ELISA kit (Invitrogen, ThermoFisher Scientific, #BMS2017) according to the manufacturer’s instructions. The standard curve was calculated from blank-curated mean standard values with a 4-parameter curve fit (R package dr4pl, v2.0.0). All sample values were blank-curated before concentration calculation via the standard curve formula.

### Data visualization

Figures were generated by R software 3.6.3, using ggplot2 package.

### Statistics

Associations between the SNP genotypes and fibroscan values or grouped fibrosis (cut-off 9.7kPa) were investigated with generalized linear models adjusting for age, BMI, gender and *PNPLA3* rs738409 with the *glm* function of the R package stats. Due to its potential as a genetic risk factor for MASLD (22), we additionally adjusted for the *PNPLA3* rs738409 genotype in all SNP-based generalized linear model (glm) calculations. The *PNPLA3* rs738409 genotyping data was available for samples of our MASLD patient cohort data but were generated in a previous study (23). Statistical analysis of this data with the *glm* function confirmed primary findings from this study (Fig. 2C).

Correlations between mycobiome and clinical data were assessed by Spearman’s correlation adjusting for age, gender, and obesity-related parameters (age, sex, BMI, DM, aHT and hyperlipidemia) using the function *pcor.test* from R package ppcor. Differentially abundant genera were identified by the Wilcoxon rank-sum test using R package stats, and by a generalized linear model adjusting for previously mentioned parameters (genus ∼ fibroscan.group + age + gender + BMI + DM + aHT + hyperlipidemia), with *glm* function from R package stats. Association between the presence or absence of CTG species and the fibrosis state was calculated by the Fisher test, using the *fisher.test* function from R package stats. A generalized linear model adjusting for previously mentioned parameters was used to study the association between CTG species and fibroscan value (genus ∼ fibroscan + age + gender + BMI + DM + aHT + hyperlipidemia), with *glm* function from R package stats. When exploring all data, the antibiotic intake was included for adjustment when appropriate.

### Ethics approval & consent to participate

This study, involving the MASLD patient cohort (University of Würzburg: EK 96/12, 05.09.2012; EK 188/17, 13.01.2020) and healthy volunteers (University of Würzburg: EK 191/21, 16.08.2021) was approved by the local ethics committee and conforms to the ethical guidelines of the 1975 Declaration of Helsinki. We obtained written informed consent from all patients and healthy volunteers included in this study.

## Data availability

Raw sequences from ITS1 gene sequencing were registered at NCBI under BioProject PRJNA834619.

## AUTHOR CONTRIBUTIONS

O.K., A.G., T.D. and G.P. conceived and designed the study. A.G., H.M.H., and M.R. recruited the participants and were responsible for clinical data collection. M.R. and H.M.H. collected fecal samples and extracted DNA from feces together with N.T.. A.M.A. and M.H. collected blood samples for the T cell stimulation assay, J.L. provided additional samples for this assay. H.M.H. and N.T. extracted DNA from blood and PBMC samples. R.M., N.E.N. and A.M.A. were involved in planning of experimental analyses. N.T. performed and analyzed the experimental analyses. G.P., S.L.S., O.K. and M.H. were involved in planning of mycobiome analysis. S.L.S. and M.M. performed the metagenomics analyses. K.H. and A.Sh. performed the Luminex assays. A.Sch., N.R. and I.S.B. contributed to functional *ex vivo* assays. O.K., G.P., and A.G. led and supervised the research work. N.T. and S.L.S. wrote the manuscript. O.K., A.G., G.P., R.M., K.H. and M.R. edited the manuscript. All authors reviewed and made substantial contributions and approved the final version of the manuscript.

## ACKNOWLEDGMENTS

This project was funded by the IZKF Würzburg (project A-401), the Marie Sklodowska-Curie Actions (MSCA), and Innovative Training Networks, H2020-MSCA-ITN-2018, 813781 “BestTreat” and by funds from the Deutsche Forschungsgemeinschaft (DFG) within the Collaborative Research Center CRC 124 FungiNet (project C3 to O. Kurzai; project B2 to T. Dandekar). GP would like to thank the Deutsche Forschungsgemeinschaft (DFG, German Research Foundation) under Germany’s Excellence Strategy and the Federal Ministry of Education and Research (BMBF, Germany), under the project PerMiCCion (Project ID 01KD2101A). We want to thank Ina Gaube, Barbara Conrad, Carolin Spielau-Romer and Lea Strobel for their contribution to this project.

## ABBREVIATIONS

aHT: arterial hypertension
ALD: alcohol-associated liver disease
ALT: alanine aminotransferase
AST: aspartate aminotransferase
BCAA: branched-chain amino acid
BS: bland steatosis
*C. albicans*: *Candida albicans*
*D. hansenii*: *Debaryomyces hansenii*
glm: generalized linear model
HC: healthy control
MAF: minor allele frequency
MASLD: metabolic dysfunction-associated steatotic liver disease
MASH: metabolic dysfunction-associated steatohepatitis
OTU: operational taxonomic unit
TE: transient elastography
rTreg: resting regulatory T cells
*S. cerevisiae*: *Saccharomyces cerevisiae*
SCFA: short-chain fatty acid

## REFERENCES

1. Rinella ME, Lazarus JV, Ratziu V, Francque SM, Sanyal AJ, Kanwal F, et al. A multisociety Delphi consensus statement on new fatty liver disease nomenclature. J Hepatol. 2023;79(6):1542–56.

2. Younossi Z, Anstee QM, Marietti M, Hardy T, Henry L, Eslam M, et al. Global burden of NAFLD and NASH: trends, predictions, risk factors and prevention. Nat Rev Gastroenterol Hepatol. 2018;15(1):11–20.

3. Friedman SL, Neuschwander-Tetri BA, Rinella M, and Sanyal AJ. Mechanisms of NAFLD development and therapeutic strategies. Nat Med. 2018;24(7):908–22.

4. Donnelly KL, Smith CI, Schwarzenberg SJ, Jessurun J, Boldt MD, and Parks EJ. Sources of fatty acids stored in liver and secreted via lipoproteins in patients with nonalcoholic fatty liver disease. J Clin Invest. 2005;115(5):1343–51.

5. Buzzetti E, Pinzani M, and Tsochatzis EA. The multiple-hit pathogenesis of non-alcoholic fatty liver disease (NAFLD). Metabolism. 2016;65(8):1038–48.

6. Rau M, Schilling AK, Meertens J, Hering I, Weiss J, Jurowich C, et al. Progression from Nonalcoholic Fatty Liver to Nonalcoholic Steatohepatitis Is Marked by a Higher Frequency of Th17 Cells in the Liver and an Increased Th17/Resting Regulatory T Cell Ratio in Peripheral Blood and in the Liver. J Immunol. 2016;196(1):97–105.

7. Tripathi A, Debelius J, Brenner DA, Karin M, Loomba R, Schnabl B, et al. The gut-liver axis and the intersection with the microbiome. Nat Rev Gastroenterol Hepatol. 2018;15(7):397–411.

8. Sharma S, and Tripathi P. Gut microbiome and type 2 diabetes: where we are and where to go? J Nutr Biochem. 2019;63:101–8.

9. Palmas V, Pisanu S, Madau V, Casula E, Deledda A, Cusano R, et al. Gut microbiota markers associated with obesity and overweight in Italian adults. Sci Rep. 2021;11(1):5532.

10. Lang S, and Schnabl B. Microbiota and Fatty Liver Disease-the Known, the Unknown, and the Future. Cell Host Microbe. 2020;28(2):233–44.

11. Sharpton SR, Schnabl B, Knight R, and Loomba R. Current Concepts, Opportunities, and Challenges of Gut Microbiome-Based Personalized Medicine in Nonalcoholic Fatty Liver Disease. Cell Metab. 2021;33(1):21–32.

12. Leung H, Long X, Ni Y, Qian L, Nychas E, Siliceo SL, et al. Risk assessment with gut microbiome and metabolite markers in NAFLD development. Sci Transl Med. 2022;14(648):eabk0855.

13. Liu Y, Meric G, Havulinna AS, Teo SM, Aberg F, Ruuskanen M, et al. Early prediction of incident liver disease using conventional risk factors and gut-microbiome-augmented gradient boosting. Cell Metab. 2022;34(5):719–30 e4.

14. Rau M, Rehman A, Dittrich M, Groen AK, Hermanns HM, Seyfried F, et al. Fecal SCFAs and SCFA-producing bacteria in gut microbiome of human NAFLD as a putative link to systemic T-cell activation and advanced disease. United European Gastroenterol J. 2018;6(10):1496–507.

15. Nash AK, Auchtung TA, Wong MC, Smith DP, Gesell JR, Ross MC, et al. The gut mycobiome of the Human Microbiome Project healthy cohort. Microbiome. 2017;5(1):153.

16. Thielemann N, Herz M, Kurzai O, and Martin R. Analyzing the human gut mycobiome - A short guide for beginners. Comput Struct Biotechnol J. 2022;20:608–14.

17. Turner SA, and Butler G. The Candida pathogenic species complex. Cold Spring Harb Perspect Med. 2014;4(9):a019778.

18. Bacher P, Hohnstein T, Beerbaum E, Rocker M, Blango MG, Kaufmann S, et al. Human Anti-fungal Th17 Immunity and Pathology Rely on Cross-Reactivity against Candida albicans. Cell. 2019;176(6):1340–55 e15.

19. Demir M, Lang S, Hartmann P, Duan Y, Martin A, Miyamoto Y, et al. The fecal mycobiome in non-alcoholic fatty liver disease. J Hepatol. 2022;76(4):788–99.

20. Zeng S, Rosati E, Saggau C, Messner B, Chu H, Duan Y, et al. Candida albicans-specific Th17 cell-mediated response contributes to alcohol-associated liver disease. Cell Host Microbe. 2023;31(3):389–404 e7.

21. Arslanow A, Stokes CS, Weber SN, Grunhage F, Lammert F, and Krawczyk M. The common PNPLA3 variant p.I148M is associated with liver fat contents as quantified by controlled attenuation parameter (CAP). Liver Int. 2016;36(3):418–26.

22. Salari N, Darvishi N, Mansouri K, Ghasemi H, Hosseinian-Far M, Darvishi F, et al. Association between PNPLA3 rs738409 polymorphism and nonalcoholic fatty liver disease: a systematic review and meta-analysis. BMC Endocr Disord. 2021;21(1):125.

23. Sarlos P, Kovesdi E, Magyari L, Banfai Z, Szabo A, Javorhazy A, et al. Genetic update on inflammatory factors in ulcerative colitis: Review of the current literature. World J Gastrointest Pathophysiol. 2014;5(3):304–21.

24. Borman AM, and Johnson EM. Name Changes for Fungi of Medical Importance, 2018 to 2019. J Clin Microbiol. 2021;59(2).

25. Seelbinder B, Chen J, Brunke S, Vazquez-Uribe R, Santhaman R, Meyer AC, et al. Antibiotics create a shift from mutualism to competition in human gut communities with a longer-lasting impact on fungi than bacteria. Microbiome. 2020;8(1):133.

26. Watts SC, Ritchie SC, Inouye M, and Holt KE. FastSpar: rapid and scalable correlation estimation for compositional data. Bioinformatics. 2019;35(6):1064–6.

27. Eddowes PJ, Sasso M, Allison M, Tsochatzis E, Anstee QM, Sheridan D, et al. Accuracy of FibroScan Controlled Attenuation Parameter and Liver Stiffness Measurement in Assessing Steatosis and Fibrosis in Patients With Nonalcoholic Fatty Liver Disease. Gastroenterology. 2019;156(6):1717–30.

28. Butler G, Rasmussen MD, Lin MF, Santos MA, Sakthikumar S, Munro CA, et al. Evolution of pathogenicity and sexual reproduction in eight Candida genomes. Nature. 2009;459(7247):657-62.

29. Ramos-Moreno L, Ruiz-Perez F, Rodriguez-Castro E, and Ramos J. Debaryomyces hansenii Is a Real Tool to Improve a Diversity of Characteristics in Sausages and Dry-Meat Products. Microorganisms. 2021;9(7).

30. Loomba R, Friedman SL, and Shulman GI. Mechanisms and disease consequences of nonalcoholic fatty liver disease. Cell. 2021;184(10):2537–64.

31. McGeachy MJ, Cua DJ, and Gaffen SL. The IL-17 Family of Cytokines in Health and Disease. Immunity. 2019;50(4):892–906.

32. Conti HR, and Gaffen SL. IL-17-Mediated Immunity to the Opportunistic Fungal Pathogen Candida albicans. J Immunol. 2015;195(3):780–8.

33. Ramani K, and Biswas PS. Interleukin-17: Friend or foe in organ fibrosis. Cytokine. 2019;120:282–8.

34. Sokol H, Leducq V, Aschard H, Pham HP, Jegou S, Landman C, et al. Fungal microbiota dysbiosis in IBD. Gut. 2017;66(6):1039–48.

35. Hartmann P, Lang S, Zeng S, Duan Y, Zhang X, Wang Y, et al. Dynamic Changes of the Fungal Microbiome in Alcohol Use Disorder. Front Physiol. 2021;12:699253.

36. Shao TY, Ang WXG, Jiang TT, Huang FS, Andersen H, Kinder JM, et al. Commensal Candida albicans Positively Calibrates Systemic Th17 Immunological Responses. Cell Host Microbe. 2019;25(3):404–17 e6.

37. Chu H, Duan Y, Lang S, Jiang L, Wang Y, Llorente C, et al. The Candida albicans exotoxin candidalysin promotes alcohol-associated liver disease. J Hepatol. 2020;72(3):391–400.

38. Li XV, Leonardi I, Putzel GG, Semon A, Fiers WD, Kusakabe T, et al. Immune regulation by fungal strain diversity in inflammatory bowel disease. Nature. 2022;603(7902):672-8.

39. Brunke S, and Hube B. Two unlike cousins: Candida albicans and C. glabrata infection strategies. Cell Microbiol. 2013;15(5):701–8.

40. Ochangco HS, Gamero A, Smith IM, Christensen JE, Jespersen L, and Arneborg N. In vitro investigation of Debaryomyces hansenii strains for potential probiotic properties. World J Microbiol Biotechnol. 2016;32(9):141.

41. Jain U, Ver Heul AM, Xiong S, Gregory MH, Demers EG, Kern JT, et al. Debaryomyces is enriched in Crohn’s disease intestinal tissue and impairs healing in mice. Science. 2021;371(6534):1154-9.

42. Frau A, Kenny JG, Lenzi L, Campbell BJ, Ijaz UZ, Duckworth CA, et al. DNA extraction and amplicon production strategies deeply inf luence the outcome of gut mycobiome studies. Sci Rep. 2019;9(1):9328.

43. Chu H, Duan Y, Yang L, and Schnabl B. Small metabolites, possible big changes: a microbiota-centered view of non-alcoholic fatty liver disease. Gut. 2019;68(2):359–70.

44. Guinan J, Wang S, Hazbun TR, Yadav H, and Thangamani S. Antibiotic-induced decreases in the levels of microbial-derived short-chain fatty acids correlate with increased gastrointestinal colonization of Candida albicans. Sci Rep. 2019;9(1):8872.

45. Tovo CV, Villela-Nogueira CA, Leite NC, Panke CL, Port GZ, Fernandes S, et al. Transient hepatic elastography has the best performance to evaluate liver fibrosis in non-alcoholic fatty liver disease (NAFLD). Ann Hepatol. 2019;18(3):445–9.

46. European Association for Study of L, and Asociacion Latinoamericana para el Estudio del H. EASL-ALEH Clinical Practice Guidelines: Non-invasive tests for evaluation of liver disease severity and prognosis. J Hepatol. 2015;63(1):237–64.

47. Gweon HS, Oliver A, Taylor J, Booth T, Gibbs M, Read DS, et al. PIPITS: an automated pipeline for analyses of fungal internal transcribed spacer sequences from the Illumina sequencing platform. Methods Ecol Evol. 2015;6(8):973–80.

48. Caporaso JG, Kuczynski J, Stombaugh J, Bittinger K, Bushman FD, Costello EK, et al. QIIME allows analysis of high-throughput community sequencing data. Nat Methods. 2010;7(5):335–6.

49. Nilsson RH, Larsson KH, Taylor AFS, Bengtsson-Palme J, Jeppesen TS, Schigel D, et al. The UNITE database for molecular identification of fungi: handling dark taxa and parallel taxonomic classifications. Nucleic Acids Res. 2019;47(D1):D259–D64.

50. Quast C, Pruesse E, Yilmaz P, Gerken J, Schweer T, Yarza P, et al. The SILVA ribosomal RNA gene database project: improved data processing and web-based tools. Nucleic Acids Res. 2013;41(Database issue):D590-6.

