## Supplemental Data for "Mycobiome Dysbiosis and Genetic Predisposition for Elevated IL-17A Drive Fibrosis in MASLD"

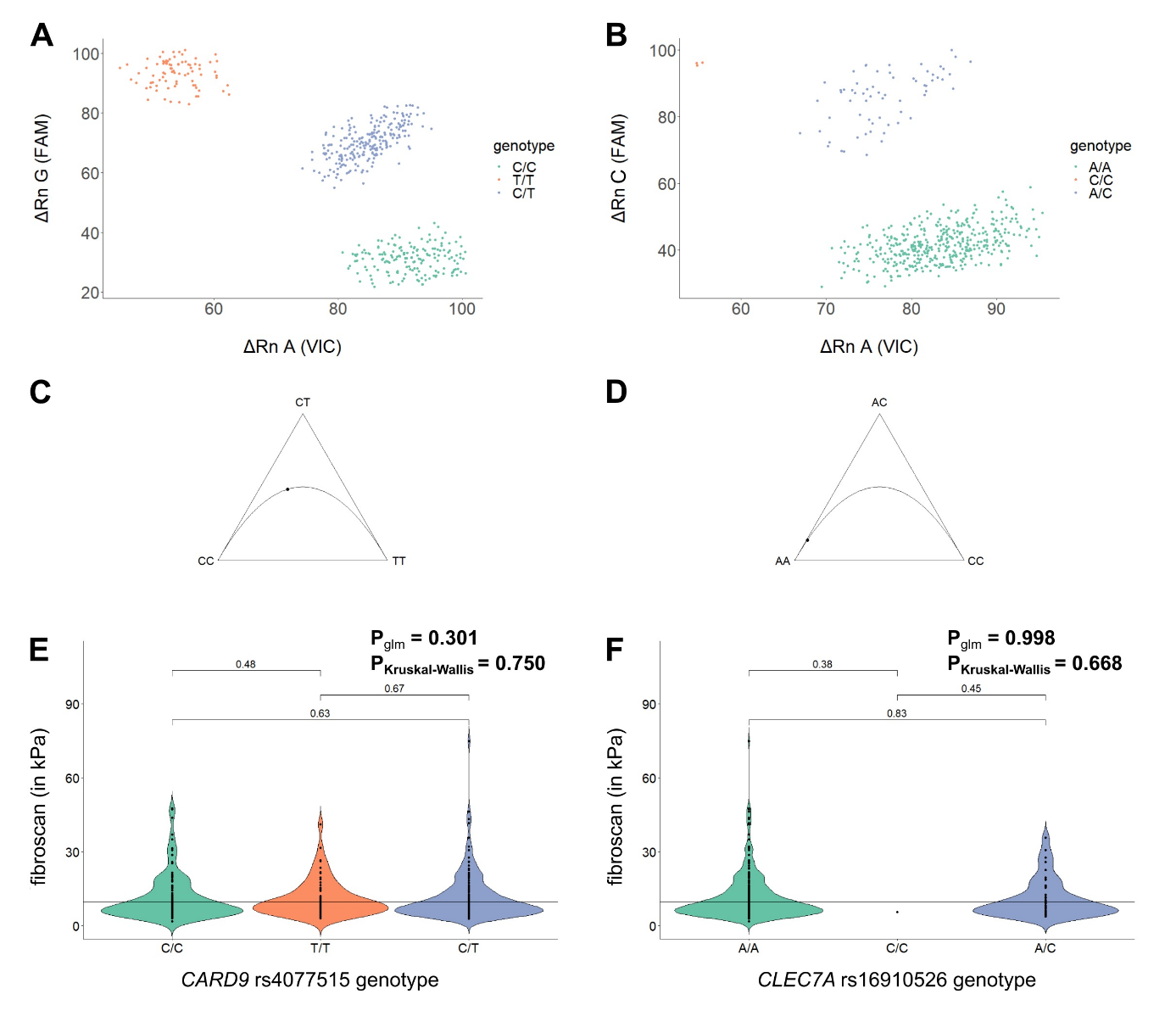

**Suppl. Fig. 1|** TaqMan SNP genotyping data for *CARD9* rs4077515 and *CLEC7A* rs16910526. Allelic discrimination plots after genotyping for *CARD9* rs4077515 **A**) and *CLEC7A* rs16910526 **B**) Ternary Plot for evaluation of Hardy-Weinberg equilibrium for *CARD9* rs4077515 **C**) and *CLEC7A* rs16910526 **D**) Violin Plot for visualization of genotype association for *CARD9* rs4077515 **E**) and *CLEC7A* rs16910526 **F**) to fibroscan values. Statistical comparisons were performed using generalized linear models adjusted for age, gender, BMI, *PNPLA3* rs738409 genotype based on a fibroscan cut-off=9.7 kPa but were not significant (p_glm_ (rs4077515)=0.3, p_glm_(rs16910526)=0.5).

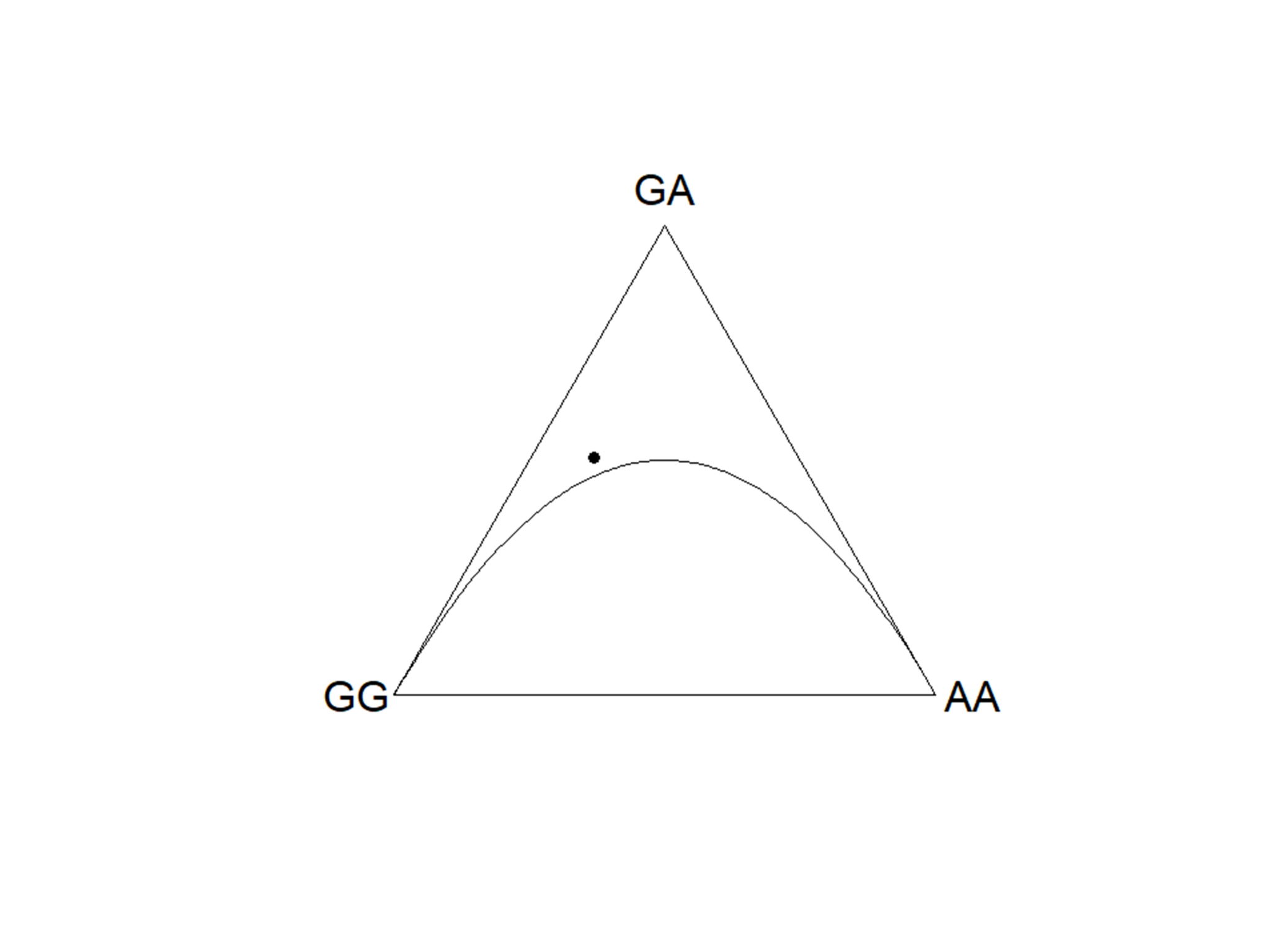

**Suppl. Fig. 2|** Ternary Plot of *IL17A* rs2275913 data. *IL17A* rs2275913 genotyping data are in Hardy-Weinberg equilibrium and thereby selection for specific genotypes was excluded.

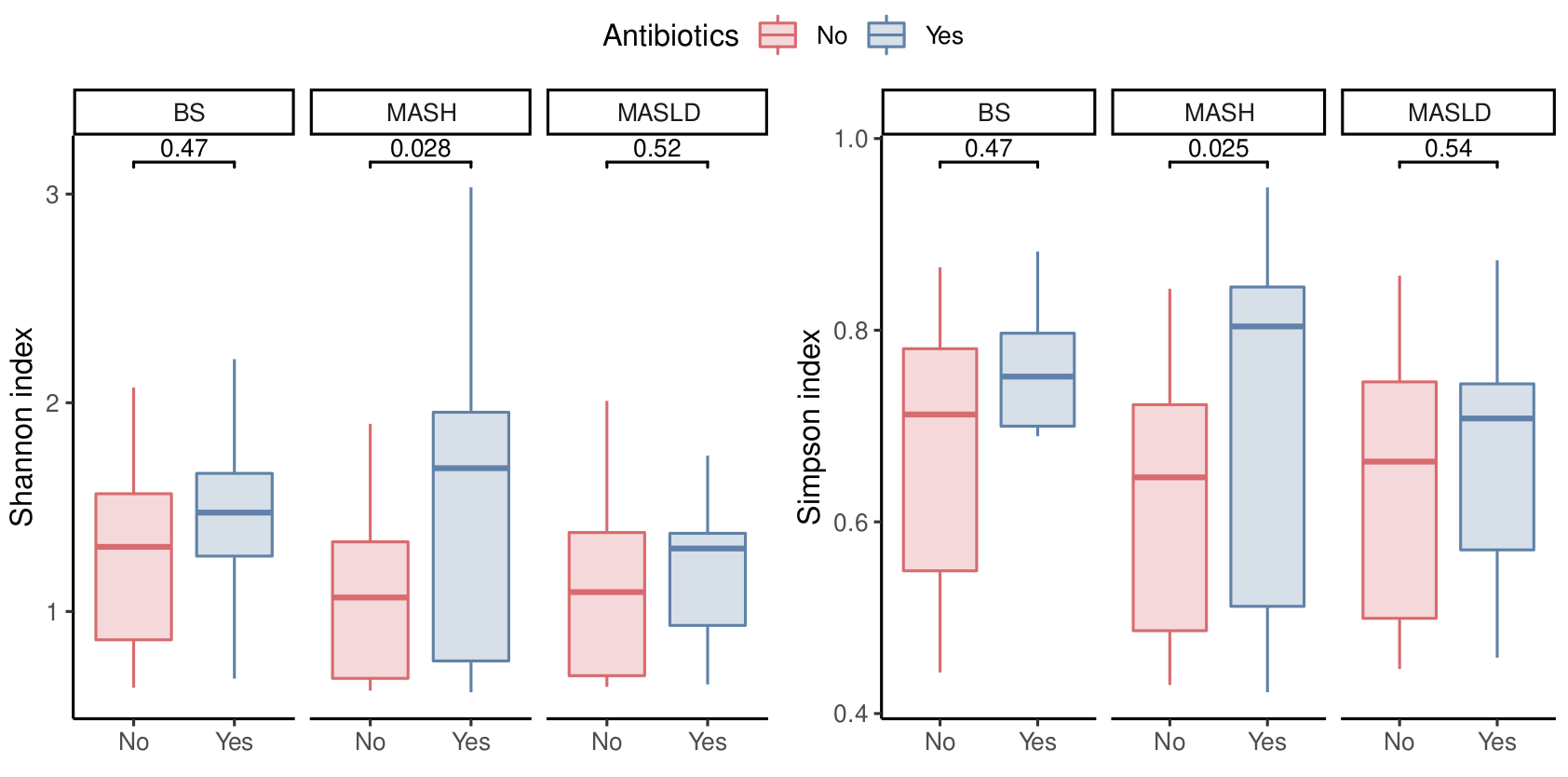

**Suppl. Fig. 3|** Comparison of Shannon (left) and Simpson (right) indexes between antibiotic-free subjects (No, red) and subjects that used antibiotics within the six months prior to the sample collection (Yes, blue) in BS, MASH and MASLD groups.

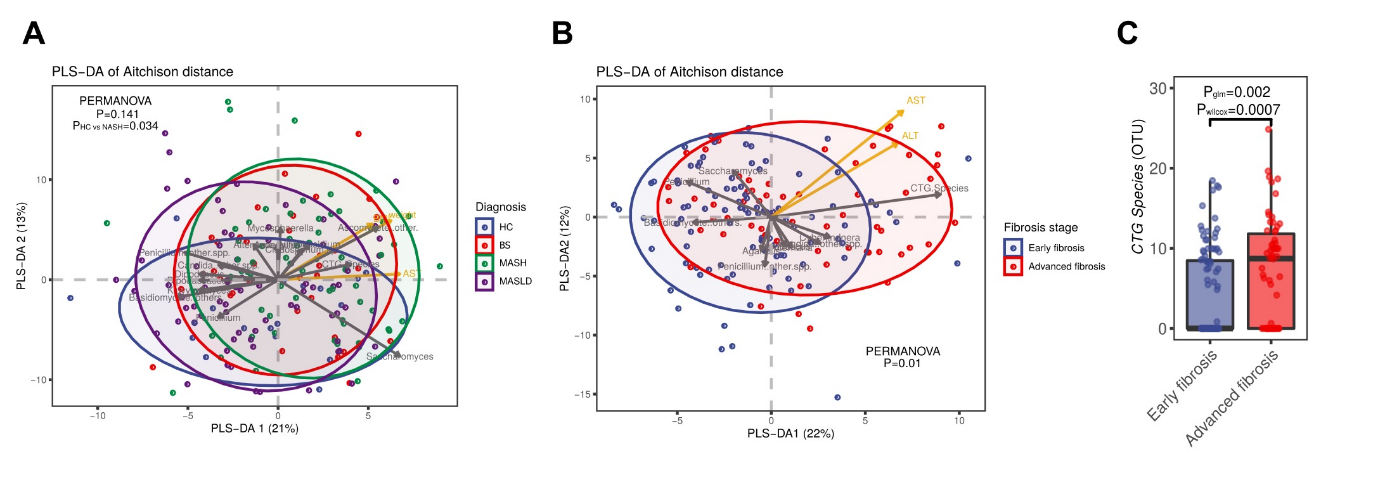

**Suppl. Fig. 4|** Mycobiome changes using the full cohort of samples. **A**) Beta diversity. PLS-DA of Aitchison distance of the mycobiome composition by diagnosis. **B**) Beta diversity. PLS-DA of Aitchison distance of the mycobiome composition by fibrosis stage group. **C**) Boxplot of CTG species abundances. Statistical comparison between early and advanced fibrosis was performed using: Wilcoxon rank-sum test (p_wilcoxon_) and generalized linear models adjusting for age, gender and obesity-related parameters and antibiotic intake (p_glm_).

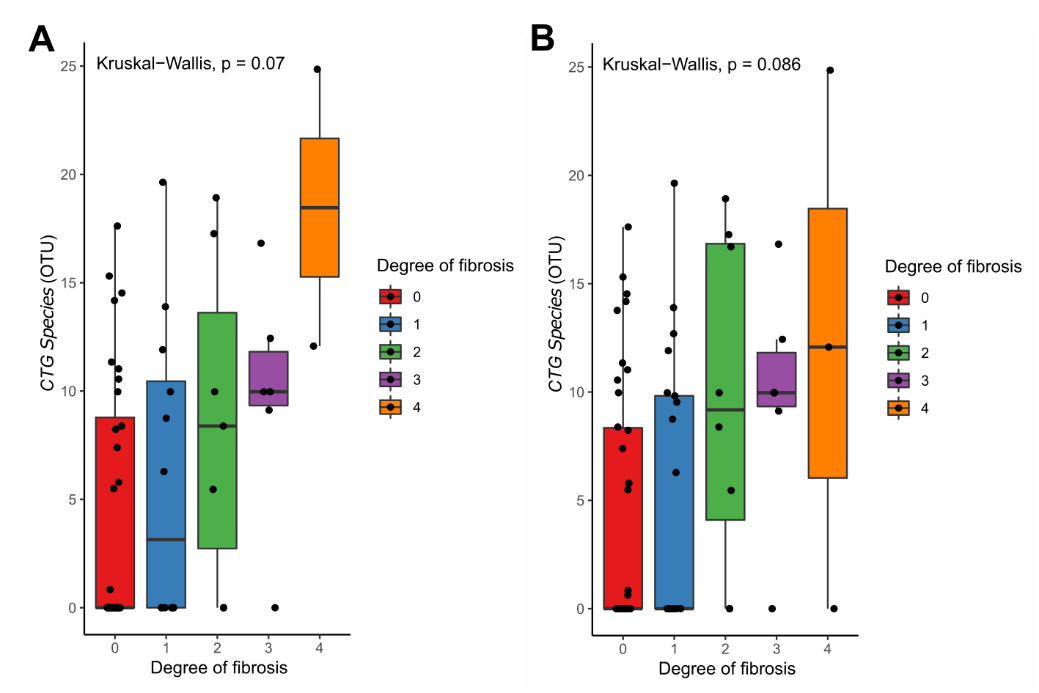

**Suppl. Fig. 5|** Boxplot of CTG species abundances. **A**) Antibiotic-free set of samples. **B**) Full cohort. Statistical comparison between fibrosis stages (obtained by biopsy) were performed using Kruskal-Wallis test.

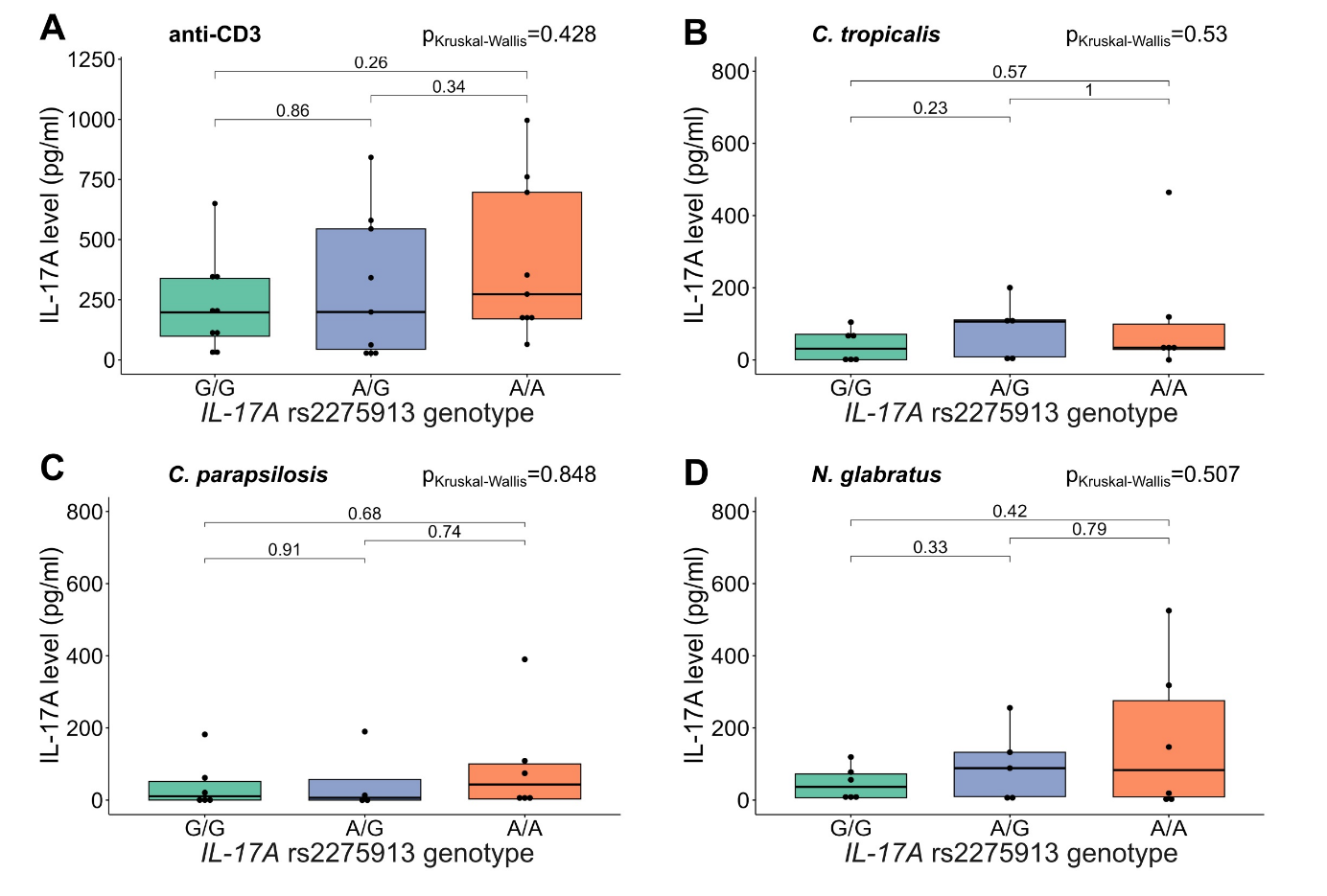

**Suppl. Fig. 6|** IL-17A secretion in T cells after stimulation with anti-CD3 as stimulation control and fungal lysates. IL-17A concentrations in samples were measured by ELISA and calculated with a 4-parameter standard fit curve. 27 subjects were included in this assay (G/G: *n=9*, A/G: *n=9*, A/A: *n=9*). Due to interindividual variation of T cell numbers not all stimuli were tested for each condition. Deviations in samples for each genotype were indicated for the corresponding plots. **A**) IL-17A secretion after stimulation with anti-CD3 as control for sufficient T cell activation in all analyzed samples. **B-D**) IL-17A secretion after stimulation with **B**) *C. tropicalis* lysate, **C**) *C. parapsilosis* lysate. and **D**) *N. glabratus* lysate. Statistical comparisons for **A**-**D** were performed using Kruskal-Wallis Test (p_Kruskal-Wallis_) and T test comparing mean IL-17A values between genotypes. Horizontal lines in the boxplots indicate from top to bottom 75th percentile, median and 25th percentile. Whiskers display minimum and maximum values in 1.5x the interquartile range. Dots specify individuals for the three *IL17A* rs2275913 genotypes.
